# Clinical and Molecular Effects of TYK2/JAK1 Inhibition in Dermatomyositis

**DOI:** 10.64898/2026.06.15.26354270

**Authors:** AR Mangold, RA Vleugels, JJ Paik, N Shahriari, RL Castillo, JG Gehlhausen, R Jiang, JC Sluzevich, A Haemel, J Fox, R Bogle, BT Roberts, S Penner, X Li, Z Ramirez, LC Tsoi, KS Shaw, MD Cascino, BM Johnson, JM Kahlenberg, L Christopher-Stine, AP Fernandez, DF Fiorentino, VP Werth, JE Gudjonsson

**Author notes:** =contributed equally as co-first authors (AR Mangold, RA Vleugels). =contributed equally as co-senior authors (VP Werth, JE Gudjonsson). **Corresponding Author:** Ruth Ann Vleugels, MD, MBA, MPH, Department of Dermatology, Mass General Brigham, Boston, MA 02115. **Funding sources:** The study was funded by Priovant Therapeutics. **IRB approval status:** The study was approved by the Mayo Clinic Institutional Review Board.

## Abstract

Dermatomyositis is driven by overactivation of type I and II interferons and other proinflammatory cytokines that signal via the JAK-STAT pathway. We conducted a 12-week, open-label study of brepocitinib, an oral TYK2/JAK1 inhibitor, in five adults with severe cutaneous dermatomyositis. Treatment was associated with rapid, clinically meaningful improvement in cutaneous disease activity. Single-cell and spatial transcriptomic profiling of lesional skin showed marked suppression of interferon-responsive pathways and inflammatory cell states by week 4. Together with findings from a Phase 3 randomized trial in DM patients with skin and muscle involvement (VALOR, NCT05437263), these data support TYK2/JAK1 inhibition as a promising therapeutic strategy for DM.

## Introduction

Dermatomyositis (DM) is a highly morbid, systemic autoimmune disease characterized by dysregulated type I and II interferon (IFN) signaling.^1,2^ IFN-stimulated genes (ISGs) are consistently overexpressed in the blood, skin, and muscle of affected patients and closely correlate with clinical disease activity across organ system domains.^3,4^ This IFN-rich inflammatory milieu recruits and sustains cytotoxic CD8+ effector T cells, activated B cells, and other immune cell populations, promoting interface dermatitis in the skin and perivascular inflammation with perifascicular atrophy and myofiber degeneration in the muscle.^5,6^

Tyrosine kinase 2 (TYK2) and Janus kinase 1 (JAK1) mediate signaling of multiple pro-inflammatory cytokines implicated in DM pathogenesis including type I and II IFNs.^7^ Thus, selective inhibition of TYK2 and JAK1 represents a promising therapeutic approach for mitigating these cytokine-driven inflammatory pathways. We conducted a single-center, open-label, Phase 2 study evaluating brepocitinib, an oral TYK2/JAK1 inhibitor, in adults with severe, cutaneous DM and performed single-cell and spatial transcriptomic profiling of lesional skin to characterize the molecular effects of targeted pathway inhibition.

## Results

### Clinical Course

Clinical and demographic data of study participants are summarized in **Table S1**. Five participants with cutaneous DM (4 female; mean [SD] age, 53 [10] years) were enrolled. All participants had failed common first- and second-line treatments for DM; three were being treated with stable doses of conventional immunosuppressants at study initiation, including 3 taking prednisone (mean [range] prednisone dose, 8.3 mg [5–10]). All participants had severe cutaneous disease,^9^ with a mean CDASI-A score of 40.2 [range, 30–46].

All participants demonstrated rapid and clinically meaningful improvement in Cutaneous Dermatomyositis Disease Area and Severity Index-Activity (CDASI-A) score and other measures of skin disease activity (**Fig. 1**). Mean change [SD] and mean percentage change [SD] in CDASI-A from baseline were -20.8 [8.4] and -51.8% [17.8%] at Week 4, and -30.6 [6.4] and -77.0% [15.8%] at Week 12, respectively. All five participants exceeded published minimal clinically important difference (MCID) thresholds for CDASI-A by week 4 (≥ 40% relative and ≥ 4-point absolute reduction), and two achieved CDASI-A scores of ≤ 5 by Week 12, indicating functional remission of skin disease.^8,9^ Parallel changes (mean [SD]) from baseline were observed across additional clinician- and patient-reported outcome measures (**Table S2)**, including Physician’s Global Assessment Visual Analog Scale (PhGA-VAS; -5.9 [0.9]), Patient Global Assessment Visual Analog Scale (PtGA-VAS; -5.1 [1.8]), Cutaneous Dermatomyositis Activity Investigator’s Global Assessment (CDA-IGA; -2.2 [0.5]), and Patient Global Assessment-Skin (PtGA-skin; -1.6 [0.6]). Patient-reported quality of life improved as well, with a mean (SD) change in Skindex-16 score of -41.3 (10.9) by Week 12, exceeding the validated 10-point MCID and reflecting clinically meaningful improvement across emotional, functional, and symptom domains.^10^

**Figure 1.**
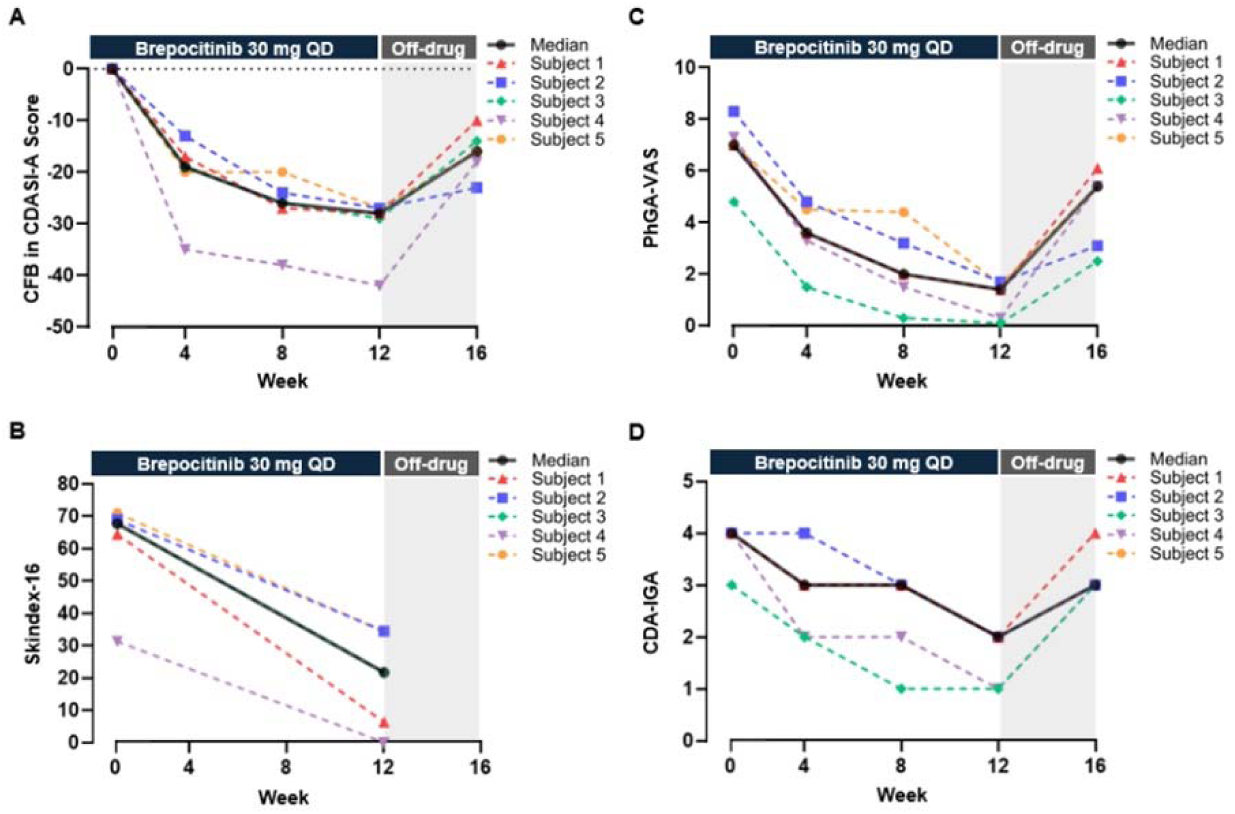
Clinical Responses to Brepocitinib in Cutaneous Dermatomyositis. Change from baseline in CDASI-A **(A)** as well as absolute values of Skindex-16 **(B)**, PhGA-VAS (**C**), and CDA-IGA **(D)** are shown for individual patients (colored dashed lines) and the group median (black solid line). Weeks 0–12 represent the treatment period with brepocitinib 30 mg once daily; weeks 12–16 represent the 4-week off-drug follow-up period. Rapid and clinically meaningful improvements were observed during the treatment period, with partial loss of response after drug discontinuation. *Abbreviations: CDASI-A, Cutaneous Dermatomyositis Disease Area and Severity Index – Activity Score; PhGA-VAS, Physician’s Global Assessment – Visual Analog Scale; CDA-IGA – Cutaneous Dermatomyositis Investigator’s Global Assessment*

Following brepocitinib discontinuation, partial loss of response was observed in all participants (mean [SD] CDASI-A score increased from 9.6 [6.7] at Week 12 to 24.0 [6.6] at Week 16), supporting a treatment-specific effect (**Fig. 1**). Brepocitinib was well-tolerated with no serious adverse events (SAEs) or discontinuations due to AEs (**Fig. S1, Table S3**).

### Molecular Responses

Single-cell and spatial RNA sequencing performed on lesional skin biopsy specimens from four participants at baseline and Week 4 yielded 100,190 high-quality cells spanning 18 major cell types (**Figs. S3, S4**).^11,12^ Immune-cell proportions, including B cells, natural killer (NK) cells, and T cells, declined rapidly after brepocitinib treatment, mirroring the kinetics of the observed clinical responses. Within the T-cell compartment, re-clustering of 6,679 cells defined nine subpopulations (**Fig. 2**), including a CD8^+^ effector/memory T-cell (Tem) subset that was abundant at baseline and enriched for early activation markers, cytotoxic mediators (*PRF1*, *GZMB*, *GZMK*), and type II IFN-associated effector programs (**Fig. S4**). By Week 4, this pathogenic subset demonstrated downregulation of key pathways essential for T-cell activation, survival, and cytotoxic function, including broad transcriptional suppression of ISGs, cytotoxic effector molecules, and IL-2/STAT5 signaling. NK cells exhibited parallel reductions in cytotoxic and IFN-responsive pathways, reflecting coordinated attenuation of effector circuits implicated in DM (**Fig. S4**).

**Figure 2.**
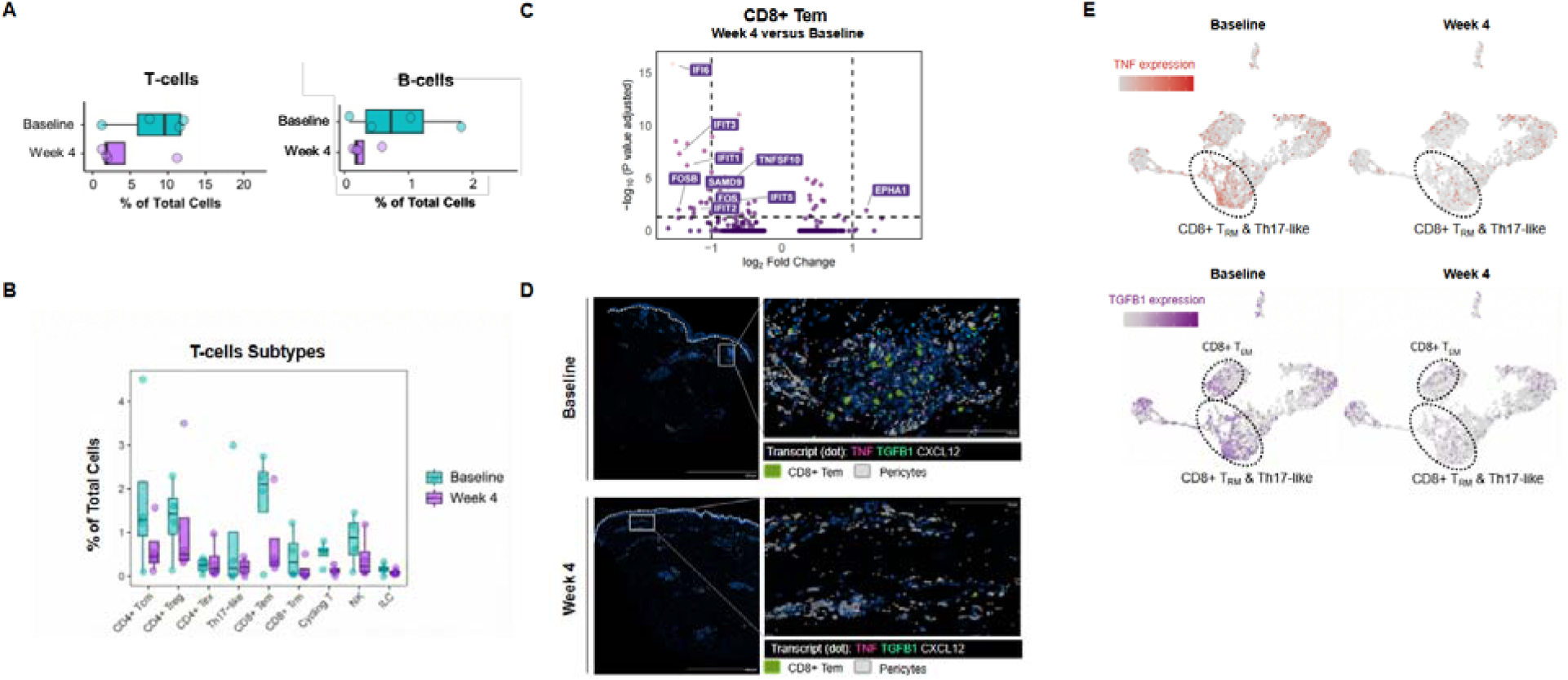
Early Cellular Responses in Cutaneous Dermatomyositis with Brepocitinib. Single-cell RNA sequencing of paired lesional dermatomyositis (DM) skin specimens demonstrated reduced proportions of activated T-cells and B-cells by week 4 of brepocitinib treatment. Boxplots (**A**) demonstrate shifts in the proportion of T-cells and B-cells out of global cells (N=4 participants), including median, first (lower hinge), and third (upper hinge) quartiles. Of the profiled lymphocyte cell populations (**B**), CD4+ central memory T cells, CD4+ regulatory T cells, CD8+ effector memory T (Tem) cells, and NK cells were enriched in lesional DM skin at baseline and were substantially reduced by week 4. Notably, differential gene expression analysis (**C**) revealed downregulation of multiple interferon-stimulated genes (ISGs) in CD8+ Tem cells. Subcellular mapping (**D**) of representative transcripts in CD8+ Tem cells and pericytes demonstrated reduced TNF/TGF-β signaling and chemokine-mediated T-cell-pericyte interactions. Feature plots (**E**) corroborated decreased expression levels of TNF and TGF-β signaling in T cell subsets.

Differential expression analyses identified keratinocytes (KCs) and pericytes as the most transcriptionally responsive cell populations (**Fig. S2**). Basal and cycling KC populations demonstrated marked reductions in canonical ISGs (IFI6, IFI27, IFIT1/2, and CXCL10/11) alongside decreased type I/II IFN module scores (**Fig. S5**). Ligand-receptor analyses revealed diminished CXCL9/11-CXCR3 interactions between KCs and CD8^+^ Tem cells, suggesting disruption of the epithelial-immune axis underlying lesional interface dermatitis in cutaneous DM.^13^ Integrated spatial transcriptomic profiling reinforced these findings, demonstrating normalization of type I and II IFN signaling at the dermal-epidermal junction by Week 4, consistent with early clinical improvement (**Fig. 3**).

**Figure 3.**
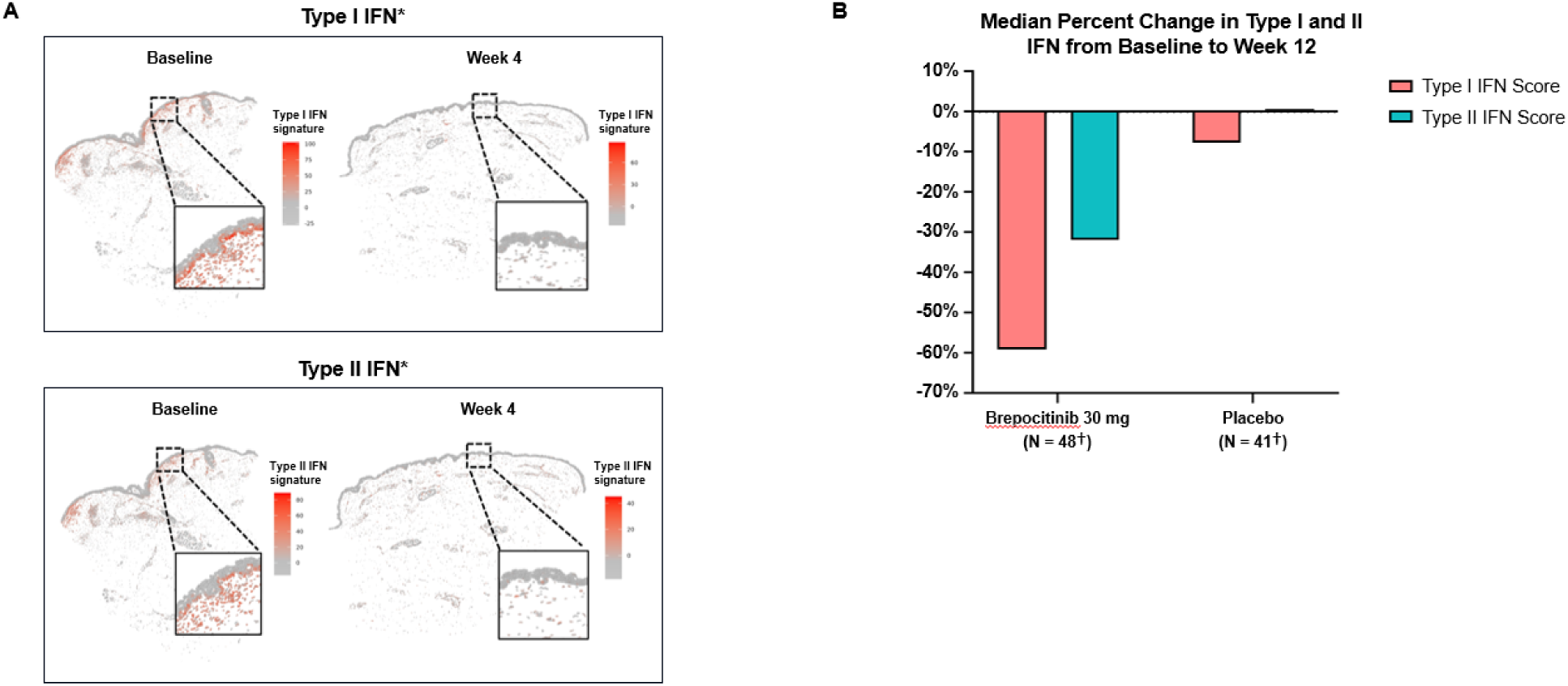
Marked Attenuation of Type I and II IFN Signaling in Dermatomyositis Skin and Blood with Brepocitinib. Spatial RNA sequencing of lesional dermatomyositis (DM) skin biopsies (**A**) obtained at baseline demonstrated prominent aggregation of type I and type II interferon (IFN) signals at the dermal-epidermal junction. By week 4 of brepocitinib treatment, marked decreases in both type I and type II IFN signaling were observed. To further contextualize these tissue-level findings, bulk RNA sequencing of whole blood from participants in a separate, concurrent, Phase 3, double-blind, randomized trial of brepocitinib in dermatomyositis (VALOR, NCT05437263) was performed (**B**). In this independent cohort, participants receiving brepocitinib 30 mg once daily demonstrated similarly rapid reductions in type I and type II IFN gene-expression scores from baseline to week 12 compared to the placebo group. *Representative spatial transcriptomic images of type I and II IFN signature scores in lesional DM skin from a single participant are shown †Number of participants with whole-blood samples available at both baseline and week 12

Pericytes also exhibited pronounced transcriptional shifts in response to brepocitinib, with broad downregulation of angiogenesis, extracellular matrix (ECM) organization, and migratory programs that are increasingly recognized as substantial contributors to microvascular injury in DM-associated myopathy.^14^ NicheNet^15^ analysis identified TNF- and TGF-β-signaling from CD8^+^ Tem cells as key upstream drivers of these changes, consistent with a model in which cytotoxic T-cell-derived cytokines activate pericytes, promote ECM remodeling, and amplify chemokine-dependent recruitment of additional immune cells (**Fig. S6**). Spatial analysis corroborated these findings, revealing dense TNF and TGF-β transcript localization within perivascular and dermal-epidermal junction regions at baseline that was markedly reduced by Week 4 of brepocitinib treatment. Concordantly, spatial co-localization of pericytes and CD8+ Tem cells decreased with brepocitinib, suggesting that brepocitinib may disrupt a self-amplifying stromal-immune circuit underlying microvascular pathology in DM (**Fig. 2**).

Notably, myeloid cell populations, important amplifiers of IFN response,^16^ demonstrated convergent suppression of IFN-driven processes (**Fig. S7**). In particular, cDC2B dendritic cells declined with treatment and exhibited transcriptional silencing of key ISGs (IFI6, IFI27, IFIT1/2, and CXCL10) and related pathways. Together, these findings underscore the collapse of the IFN-rich inflammatory microenvironment of DM across both immune and stromal compartments by Week 4 of brepocitinib treatment.

To further contextualize these tissue-level findings, bulk RNA sequencing was performed on whole-blood obtained from DM patients receiving brepocitinib 30 mg once daily in a concurrent Phase 3 study (VALOR; NCT05437263). This analysis demonstrated similarly rapid suppression of type I and II IFN gene-expression scores from baseline through Week 12 (**Fig. 3**). This parallel reduction across blood and skin reveals a consistent pharmacodynamic signature of TYK2/JAK1 inhibition in DM and aligns with the clinical efficacy observed in both the Phase 2 and Phase 3 studies.^17^

## Discussion

Cutaneous disease is a major driver of morbidity in DM.^18^ Lesions are frequently chronic, intensely pruritic, and cosmetically disfiguring, resulting in profound impairments in quality of life. Existing therapies including systemic corticosteroids, conventional immunosuppressants, and intravenous immunoglobulin offer inconsistent benefit, are associated with substantial toxicity, and can require complex or burdensome administration regimens.^19-22^ Thus, a critical need remains for effective, targeted, and well-tolerated therapies for patients with DM.

In this open-label study, brepocitinib 30 mg once daily resulted in rapid and clinically meaningful improvements in cutaneous disease activity in patients with severe, treatment-refractory DM. All participants exceeded validated MCIDs for CDASI-A as early as Week 4, and two participants achieved functional remission of skin disease by Week 12.^8,9^ Patient-reported outcomes improved in parallel, with Skindex-16 scores demonstrating large and clinically meaningful reductions in key symptoms such as pruritus, psychosocial impact, and functional impairment. These findings are particularly notable given the disproportionately high, skin-related quality-of-life burden experienced by DM patients.^23^ In addition, both clinician and patient-reported global assessments of disease activity improved, with mean changes in PhGA-VAS and PtGA-VAS exceeding proposed MCID thresholds, underscoring the clinical relevance of these responses.^24,25^ Brepocitinib was well tolerated, with no significant adverse events identified during the 12-week treatment period.

Integrated single-cell and spatial transcriptomic analyses provide a cohesive mechanistic framework for the observed clinical responses. Treatment with brepocitinib produced broad suppression of type I and II ISG expression across multiple immune, epithelial, and stromal compartments, reflecting normalization of the IFN-rich inflammatory milieu that is a defining feature of DM. Immune cell populations, including CD8⁺ Tem cells, NK cells, and myeloid dendritic cells, declined with treatment, accompanied by downregulation of gene expression processes governing activation, cytotoxicity, and survival. CD8⁺ Tem cells and NK cells,^26,27^ which exhibit prominent cytotoxic and IFN-responsive signatures implicated in DM, showed marked attenuation of ISGs, effector molecules, and IL-2/STAT5 signaling. This pattern suggests that brepocitinib may disrupt pathogenic effector circuits that drive tissue inflammation and damage in DM.^28,29^

Epithelial and stromal compartments also demonstrated substantial transcriptional remodeling. KCs showed robust reductions in type I and II IFN signaling and diminished CXCL9/11-CXCR3 interactions with cytotoxic T cells, a key epithelial-immune axis sustaining cutaneous interface dermatitis in DM.^13^ Pericytes, which are purported mediators of DM-associated microvascular injury and ECM remodeling, exhibited downregulation of angiogenesis, ECM organization, and TNF/TGF-β-driven activation processes.^30^ Myeloid populations, particularly cDC2B dendritic cells, demonstrated silencing of ISGs and associated pathways. Collectively, these findings suggest a collapse in the IFN-rich amplification loop across epithelial, stromal, and immune cell compartments with brepocitinib. Spatial profiling lends further support to these findings, as brepocitinib normalized type I and II IFN signaling at the dermal-epidermal junction and reduced co-localization of pericytes and cytotoxic CD8⁺ T cells in perivascular regions by Week 4 of treatment.

These tissue-level observations are further supported by pharmacodynamic results from a separate, Phase 3, randomized, double-blind, placebo-controlled trial in adults with DM (VALOR, NCT05437263; N=241 participants). Whole-blood RNA sequencing demonstrated similarly rapid suppression of type I and II IFN gene signatures in a larger DM population also receiving brepocitinib 30 mg once daily.^17^ This convergence across skin and blood in independent clinical cohorts supports TYK2/JAK1-mediated cytokine signaling as a central driver of disease activity in DM and provides a mechanistic rationale for targeted inhibition with brepocitinib.

While limited by small sample size, the clinical findings from this Phase 2 study are consistent with those seen in the 52-week, double-blind, placebo-controlled Phase 3 VALOR trial, the largest DM interventional trial conducted to date.^17^ Moreover, the consistency and magnitude of clinical improvement, coupled with concordant transcriptomic effects across multiple cellular compartments and their alignment with Phase 3 pharmacodynamic data, bolster the mechanistic and translational relevance of these findings in patients with DM. Together, these results provide proof-of-concept that selective TYK2/JAK1 inhibition can rapidly attenuate proinflammatory networks central to DM disease pathogenesis, supporting brepocitinib as a promising therapeutic option.

## Materials & Methods

This single-arm study (NCT06433999) enrolled five adults with severe, cutaneous DM. Participants received oral brepocitinib 30 mg once daily for 12 weeks, followed by a 4-week off-treatment monitoring period. The primary efficacy end point was the change in the CDASI-A^8^ score from baseline through Week 12. Secondary end points included clinician-reported global assessments, patient-reported outcomes, and exploratory single-cell and spatial transcriptomic analyses performed on lesional skin samples obtained at baseline and Week 4. Additional eligibility criteria and procedural details are provided in the **Supplementary Appendix**. The trial was sponsored by Priovant Therapeutics and approved by the Mayo Clinic Institutional Review Board (IRB); all participants provided written informed consent. The authors attest to the accuracy and completeness of the data.

## Conclusion

In this 12-week, open-label study of patients with severe, skin-predominant dermatomyositis, brepocitinib treatment was associated with rapid and clinically meaningful improvement in skin disease activity. Integrated single-cell and spatial transcriptomic analyses demonstrated broad suppression of interferon-driven inflammatory pathways across immune, epithelial, and stromal cell compartments, providing mechanistic insight into the observed clinical responses. These findings align with pharmacodynamic and clinical observations from the phase 3 VALOR trial and support TYK2/JAK1 inhibition as a promising, targeted therapeutic strategy for dermatomyositis.

## Data Availability

All data produced in the present work are contained in the manuscript.

