## Supplemental Appendix for "Clinical and Molecular Effects of TYK2/JAK1 Inhibition in Dermatomyositis"

**S1. Study Procedures & Methods**

**Table S1. Patient Demographics and Baseline Information**

**Table S2. Summary of Change in Clinical Efficacy Measures Through Week 12**

**Table S3. Summary of Safety**

**Figure S1. Participant Disposition**

**Figure S2. Single-Cell Transcriptomic Profiling Of Lesional Dermatomyositis With Brepocitinib Treatment**

**Figure S3. Spatial Distribution of Key Cell Types in Lesional Dermatomyositis With Brepocitinib Treatment**

**Figure S4. Characterization of Molecular Responses to Brepocitinib in T-cells**

**Fig. S5. Characterization of Molecular Responses to Brepocitinib in Keratinocytes**

**Figure S6. Characterization of Molecular Responses to Brepocitinib in Pericytes**

**Figure S7. Characterization of Molecular Responses to Brepocitinib in Myeloid Cells**

**REFERENCES**

This supplementary material has been provided by the authors to give readers additional information about their work.

**S1. Study Procedures & Methods**

*Ethics statement*

This open-label, Phase 2 trial was conducted at a single hospital-based center in the United States (Mayo Clinic, Scottsdale, Arizona). The trial was sponsored by Priovant Therapeutics, who provided the study medication and regulatory support. The study was approved by the Institutional Review Board (IRB) at the Mayo Clinic, and all participants provided written informed consent. The study was conducted in accordance with the Good Clinical Practice guidelines of the International Council for Harmonisation and the principles of the Declaration of Helsinki. The trial was registered with clinicaltrials.gov (NCT06433999).

*Participants*

Eligible patients were adults aged 18-75 years with a diagnosis of dermatomyositis consistent with the 2017 European League Against Rheumatism (EULAR)/American College of Rheumatology (ACR) Classification Criteria for Idiopathic Inflammatory Myopathies. Patients were required to have at least moderately active cutaneous dermatomyositis (defined as CDASI-A score ≥ 14), no more than minimal muscle disease (defined as MMT-8 score > 142), and at least one unsuccessful prior systemic treatment with a standard-of-care therapy. Patients were permitted to continue corticosteroid therapy of ≤ 20 mg/day of prednisone or equivalent and/or one systemic non-steroidal immunomodulatory/immunosuppressive therapy throughout the duration of the trial, as long as the total duration of therapy exceeded 12 weeks at a stable dose prior to baseline and the dose remained stable throughout the study. Patients with a history of chronic, untreated infections, active malignancy, recent thromboembolic events, or end-stage organ involvement secondary to DM that could pose additional risk to the patient or confound study assessments were excluded from the study. The first patient enrolled on August 28, 2024 and the last patient enrolled on November 8, 2024. Please see the **Protocol** for additional inclusion and exclusion criteria.

*Procedures*

Patients were screened for eligibility up to 8 weeks prior to enrollment. Patients received oral brepocitinib 30 mg once daily for 12 weeks, followed by a 4-week off-treatment monitoring period. Lesional skin biopsies were obtained at baseline and week 4.

*Clinical Outcomes*

The primary clinical efficacy end point was change from baseline in the Cutaneous Dermatomyositis Disease Area and Severity Index-Activity (CDASI-A) score through week 12. The CDASI-A is a validated outcome measure for assessing skin disease activity in DM; scores range from 0-100, with higher scores indicating more severe skin disease activity.^1^ Skin disease was further assessed by investigator- and patient-completed outcomes including the Cutaneous Dermatomyositis Activity Investigator’s Global Assessment (CDA-IGA), an investigator-completed Likert-type scale that provides an overall impression of cutaneous DM disease activity severity;^2^ the Physician’s Global Assessment Visual Analog Scale (PhGA-VAS) and Patient Global Assessment Visual Analog Scale (PtGA-VAS), which both utilize a 10-cm VAS to assess overall DM disease activity with anchors of “no evidence of disease activity” to “extremely active or severe disease activity”;^3-4^ and the PtGA-Skin, a patient-completed, single-item, Likert-type scale that captures the patient’s subjective assessment of how their DM is affecting his/her skin at the time of evaluation. Quality-of-life was assessed using the patient-completed Skindex-16 assessment tool, a validated patient-reported instrument assessing the symptomatic, emotional, and functional impact of skin disease.^5-8^ Additionally, photographs were taken at baseline, week 4, and week 12 to visually monitor treatment-related changes.

*Statistical and computational analysis*

Descriptive statistics were used to evaluate absolute values and changes in clinical efficacy measures, comparing baseline (week 0) to end-of-treatment (week 12). Single cell transcriptomic data was processed using R with the following packages: Seurat, Harmony, clusterProfiler, GSVA, iTALK, NicheNet, DEseq2, and Giotto. Analysis of spatial transcriptomics data was performed using the 10x Genomics Xenium platform.

*Dermatologic specimens and evaluation*

A 4 mm punch biopsy of lesional skin was obtained at baseline and week 4. Post-treatment initiation biopsies were taken in the same anatomic location as the pretreatment biopsies, generally 2-3 mm from the initial biopsy scar. The tissue was placed in formalin for up to 24 hours and then transitioned to 80% isopropyl alcohol before being embedded in paraffin. Single-cell RNA sequencing was performed using the Illumina NovaSeq 6000 sequencer with the assistance of the University of Michigan Advanced Genomics Core. Spatial transcriptomics analysis was performed using the Xenium spatial transcriptomics platform (10x Genomics).

*Cell clustering and cell type annotation*

Cell clustering analysis was performed using the Seurat R package (v5.0.1) on the combined gene expression matrix.^9^ Low-quality cells, defined as those with fewer than 200 detected features or with mitochondrial gene content exceeding 10%, were excluded. Gene expression levels were normalized using the NormalizeData function with default parameters. Highly variable genes were identified via the FindVariableFeatures function, followed by data scaling and centering using ScaleData. Principal component analysis (PCA) was performed on the highly variable genes, and the top 20 principal components were used in the RunHarmony function from the Harmony package (v0.1.1) to remove potential batch effect among samples processed in different libraries.^10^ UMAP dimensional reduction was performed using the RunUMAP function. Cell clusters were determined using the FindNeighbors and FindClusters functions with a resolution of 0.5. Cluster-specific marker genes were identified using the FindAllMarkers function, and cell type annotations were assigned by mapping the cluster markers to established canonical cell type signature genes.

*Cell type sub-clustering*

Sub-clustering was conducted for the most abundant cell types using the same functions described above. Sub-clusters that were defined dominantly by mitochondrial gene expression, indicating low quality, were removed from further analysis. Subtypes were annotated by cross-referencing the marker genes of sub-clusters with canonical subtype signature genes. To investigate the characteristic differences among cell clusters, the FindAllMarkers function in the Seurat package was applied to identify differentially expressed genes (DEGs) in individual clusters using the Wilcoxon rank-sum test. Genes with adjusted *P* value < 0.05 and avg_log2FC > 1 were considered upregulated, while genes with adjusted *P* value < 0.05 and avg_log2FC < -1 were classified as downregulated. Gene Ontology enrichment analyses were conducted using the clusterProfiler package (v4.8.3).

*Integration with keratinocyte cytokine signatures*

As previously described by our group,^11^ we used RNA-seq-based keratinocyte (KC) cytokine response signatures for the following cytokines: type I IFN (10 ng/mL), type II IFN (10 ng/mL), IL-17A (10 ng/mL), or TNF (10 ng/mL). Briefly, primary human KCs from 50 donors were treated with a panel of cytokines as above for 8 hours and harvested for RNA isolation. Bulk RNA-seq was performed on the Illumina NovaSeq 6000 sequencer with the assistance of the University of Michigan Advanced Genomics Core. For each stimulation condition versus unstimulated controls, differential expression analysis was performed using DESeq2.^12^ Differentially expressed genes (DEGs; two-fold increase; false discovery rate, <0.05) were used to construct response signatures for each cytokine.

*Calculation of enrichment score*

To describe the function features of different subtypes, the gene sets of 50 hallmark pathways were obtained from the MSigDB database.^13^ The score of above-mentioned pathways was calculated using gsva function in GSVA package (V1.48.3),^14^ and AUCell_calcAUC function in AUCell package (V1.22.0)^15^.

*Cell-cell interaction inference*

Inference of receptor-ligand interactions of patient-derived scRNA dataset was performed using the iTALK package. The normalized expression values were entered into the FindLR function with default parameters and visualized by the LRPlot function.^16^ The analysis was focused on cytokine interactions between pericytes and T cell subtypes. In addition, the cellular interaction was further confirmed by NicheNet (v2.0.6) package to analyze the different LR pairs between lesional week 4 and week 0 group.^17^

*Spatial sequencing data analysis*

After the Xenium run, H&E staining was performed on the Xenium slide. For quality control of Xenium analysis, the output summary HTML file confirmed that the number of detected transcripts was compatible with that in reported literature by 10x Genomics.^18^ During preprocessing, cells with fewer than 20 counts and negative features (detected in less than 5 cells) were removed from analysis. With the R package Giotto v4.3.0,^19^ a Giotto object was created based on the flat files, including cell_boundaries.csv.gz, cell_feature_matrix, etc. Information including the gene expression matrix, cell boundaries, and spatial locations of transcripts for each field of view (FOV), were incorporated into a Giotto object with the Giotto function “createGiottoObject.” Cell type annotations were performed using the Giotto function “deconvolution_results,” utilizing a reference profile from our single-cell RNA sequencing dataset. The Xenium Explorer was used to perform the gene expression analysis and define co-localization of specific cell types.

*Generation of IFN signature scores from whole blood in the Phase 3 VALOR trial*

To complement the single-cell and spatial transcriptomic analyses generated from skin specimens in this open-label, Phase 2 cohort, we also generated pharmacodynamic data from whole blood obtained during a concurrent, Phase 3, randomized, double-blind, placebo-controlled study of brepocitinib in DM (VALOR, NCT05437263). This analysis was designed to evaluate the systemic effect of brepocitinib on DM-relevant proinflammatory cytokines (specifically, Type I and II IFNs).

Bulk ribonucleic acid (RNA) sequencing was performed on whole blood samples collected at baseline and week 12 from participants in VALOR. Total RNA was extracted using standard protocols, and bulk RNA sequencing was performed to a depth of approximately 50 million paired end reads per sample. Differential expression analysis identified genes with adjusted *p* values < 0.05 and absolute log2 fold changes > 1 as significantly altered for comparisons of interest. Gene Ontology (GO) enrichment analysis was subsequently applied to the significant gene sets.

To derive interferon pathway activity metrics, a Type I IFN signature was defined using the top 20 upregulated genes annotated in the GO:0060337~type I interferon signaling pathway, while the Type II IFN signature comprised all upregulated genes in the GO:0060333~type II interferon signaling pathway. These signatures were determined using pooled baseline samples from VALOR participants compared with healthy controls and were subsequently used to quantify pharmacodynamic modulation of IFN signaling following brepocitinib treatment.

**Table S1: Patient Demographics and Baseline Information**

|  | **Patient 1** | **Patient 2** | **Patient 3** | **Patient 4** | **Patient 5** |
| --- | --- | --- | --- | --- | --- |
| Age range (years) | 51-55 | 66-70 | 41-45 | 46-50 | 51-55 |
| Sex | F | F | F | M | F |
| Disease duration (years) | < 3 | < 3 | ≥ 3 | < 3 | ≥ 3 |
| Prior therapies | TCS, SCS, MMF | TCS, SCS | HCQ, SCS, MTX, RTX, IVIg | TCS, SCS | HCQ, TCS, TCI, SCS, IVIg |
| Therapies continued with brepocitinib* | P**, MMF | P**, HCQ | None | P***, MMF | None |
| Baseline CDASI-A | 41 | 41 | 30 | 46 | 43 |

*****Permitted concomitant therapy at study entry included a stable dose of oral corticosteroids (≤ 20 mg/day of prednisone or equivalent), one antimalarial, and/or one systemic non-steroidal immunosuppressive therapy; changes in background therapy were not permitted during the 12-week treatment period

**10mg oral prednisone daily

***5mg oral prednisone daily

**Abbreviations:** DM, dermatomyositis; F, female; M, male; MTX, methotrexate; SCS, systemic corticosteroids (including prednisone, methylprednisolone); MMF, mycophenolate mofetil; TCS, topical corticosteroids; TCI, topical calcineurin inhibitors; RTX, rituximab; HCQ, hydroxychloroquine; IVIg, intravenous immune globulin; CDASI-A, Cutaneous Dermatomyositis Disease Area and Severity-Index; P, prednisone

**Table S2: Summary of Change in Clinical Efficacy Measures Through Week 12**

| Endpoint | Mean (SD) | Median (min, max) |
| --- | --- | --- |
| **CDASI-A** |  |  |
| Baseline | 40.2 (6.06) | 41.0 (30, 46) |
| CFB | -30.6 (6.43) | -28.0 (-42, -27) |
| Percent CFB | -77.0% (15.76) | -68.3% (-97, -63) |
| **CDA-IGA** |  |  |
| Baseline | 3.6 (0.89) | 4.0 (2, 4) |
| CFB | -2.0 (0.71) | -2.0 (-3, -1) |
| Percent CFB | -55.0% (11.18) | -50.0% (-75.0, -50.0) |
| **PhGA-VAS** |  |  |
| Baseline | 6.9 (1.28) | 7.0 (4.8, 8.3) |
| CFB | -6.0 (1.05) | -5.6 (-7.1, -4.7) |
| Percent CFB | -87.3% (9.30) | -85.5% (-97.9, -77.1) |
| **PtGA-VAS** |  |  |
| Baseline | 7.1 (2.64) | 7.4 (3.2, 10.0) |
| CFB | -5.1 (1.76) | -4.9 (-7.1, -2.4) |
| Percent CFB | -73.0% (13.67) | -78.7% (-82.4, -49.0) |
| **PtGA-Skin** |  |  |
| Baseline | 4.0 (0.71) | 4.0 (3, 5) |
| CFB | -1.6 (0.55) | -2.0 (-2, -1) |
| Percent CFB | -39.7% (10.83) | -40.0% (-50.0, -25.0) |
| **Skindex-16** |  |  |
| Baseline | 60.6 (16.58) | 67.7 (31.3, 71.0) |
| CFB | -41.3 (10.83) | -36.7 (-58.0, -31.3) |
| Percent CFB | -71.9% (22.50) | -67.9% (-100.0, -49.9) |

**Abbreviations**: CDA-IGA = cutaneous dermatomyositis activity investigators global assessment; CFB = change from baseline; max = maximum; min = minimum; PhGA = Physician Global Assessment; PtGA = Patient Global Assessment; SD = standard deviation; VAS = Visual Analogue Scale

**Table S3: Summary of Safety**

|  | N participants (%) [No. events] |
| --- | --- |
| All AEs | 5 (100%) [11] |
| TEAEs | 2 (40.0%) [4] |
| Serious TEAEs | 0 |
| Adverse events of special interest* | 0 |

**Abbreviations:** n, number of participants; AE, adverse event; TEAE, treatment emergent adverse event

*Adverse events of special interest were prespecified in the trial protocol and were based on brepocitinib findings across all indications and safety concerns reported for other Janus kinase and tyrosine kinase inhibitors

Two patients experienced mild adverse events (AEs) of headache and acne, which were considered treatment-related by the investigator; these were the only AEs affecting > 1 individual. There were no serious AEs, serious TEAEs, adverse events of special interest, or discontinuations due to AEs.

**Figure S1: Participant Disposition**

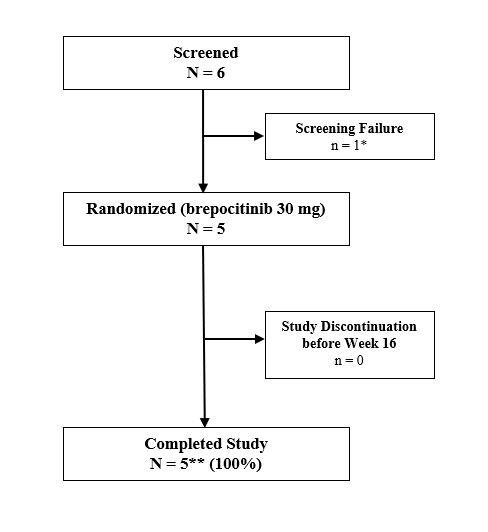

*Screen failure due to exclusion criteria

**ITT analysis set includes all enrolled participants who received at least 1 dose of brepocitinib; this was the primary analysis set for all analyses.

**Abbreviations:** ITT = intent-to-treat

**Figure S2: Single-cell Transcriptomic Profiling of Lesional Dermatomyositis Skin With Brepocitinib Treatment**

A. UMAP plot showing 100,190 cells isolated from lesional dermatomyositis (DM) skin at week 0 and week 4 of brepocitinib treatment, colored by annotated cell type.
B. UMAP visualization of treatment-associated transcriptional changes, demonstrating the number of differentially expressed genes (DEGs) per cell type at week 4 compared with baseline. Pericytes and keratinocytes exhibited the most pronounced transcriptional changes at week 4 of brepocitinib treatment.

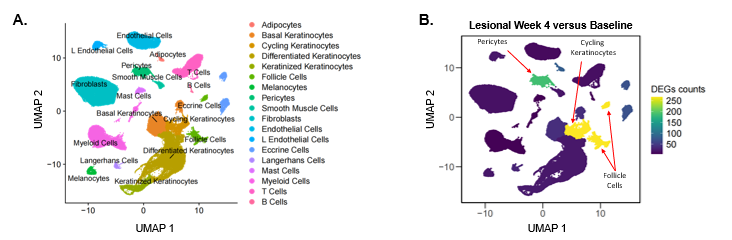

**Figure S3: Spatial Distribution of Key Cell Types in Lesional Dermatomyositis With Brepocitinib Treatment**

Annotated spatial transcriptomic maps of lesional skin biopsies from four individual patients are shown at baseline (**A**) and week 4 (**B**). Cellular identities were assigned using integrated single-cell reference profiles, enabling visualization of the spatial organization of immune, stromal, and epithelial populations across full-thickness skin sections.

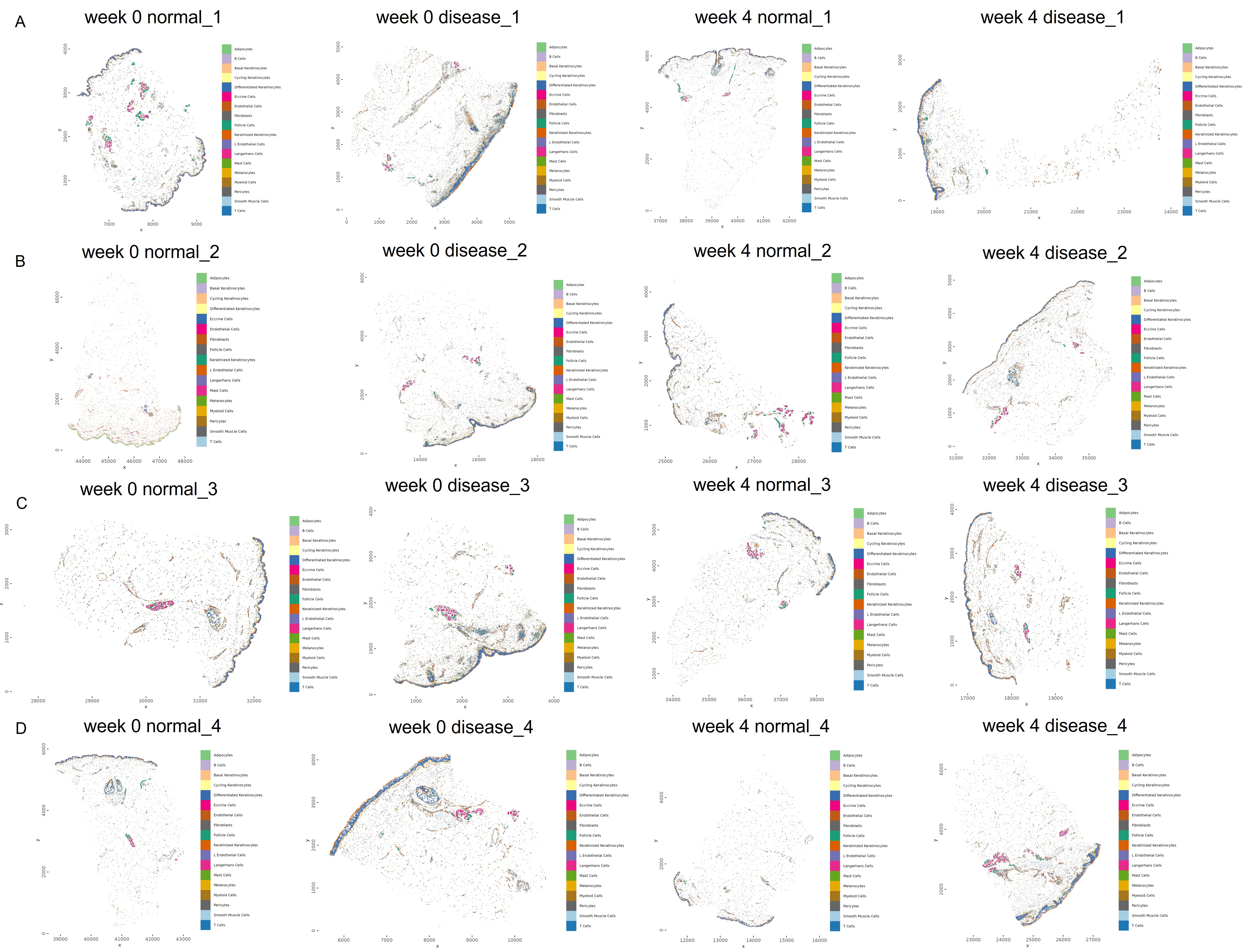

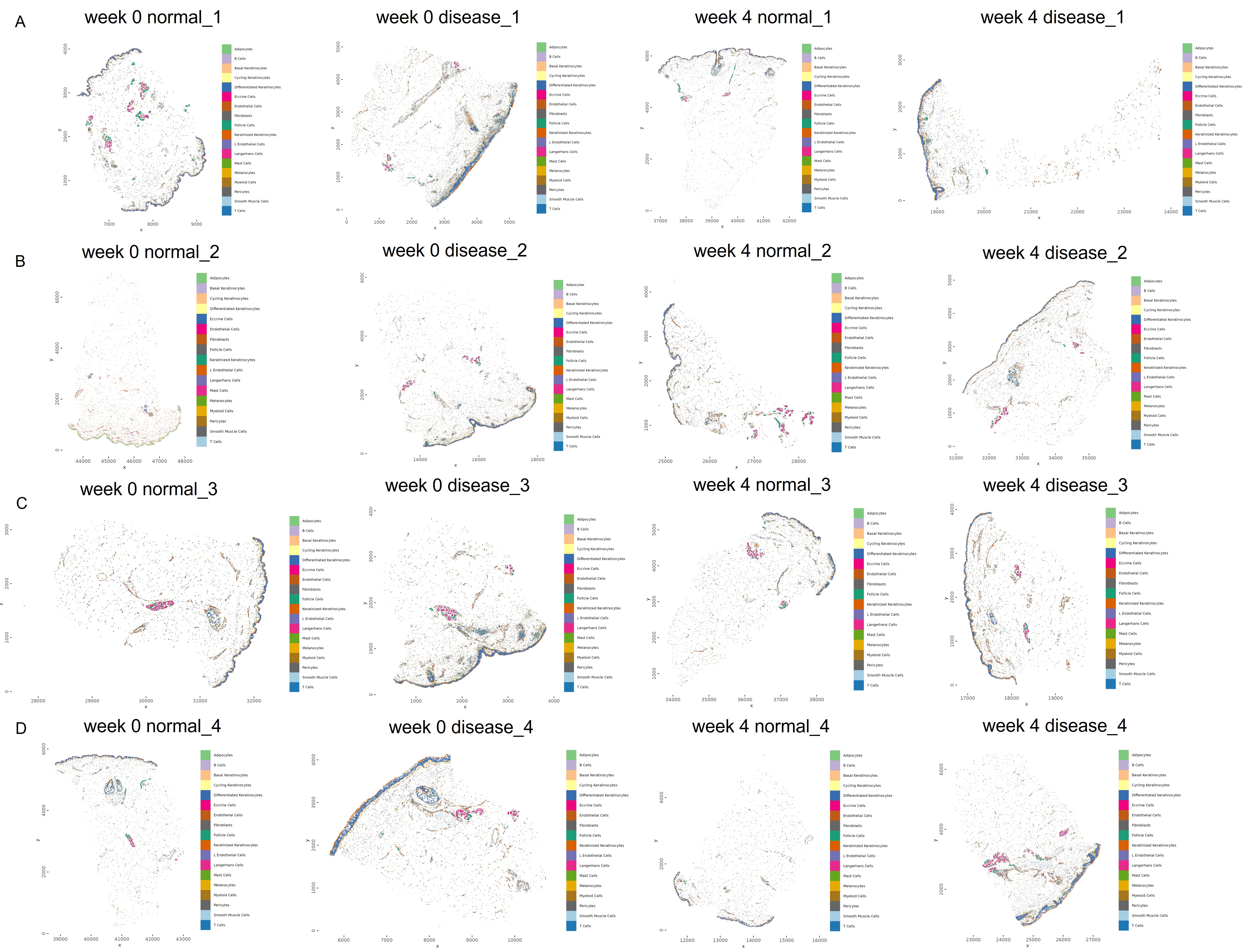

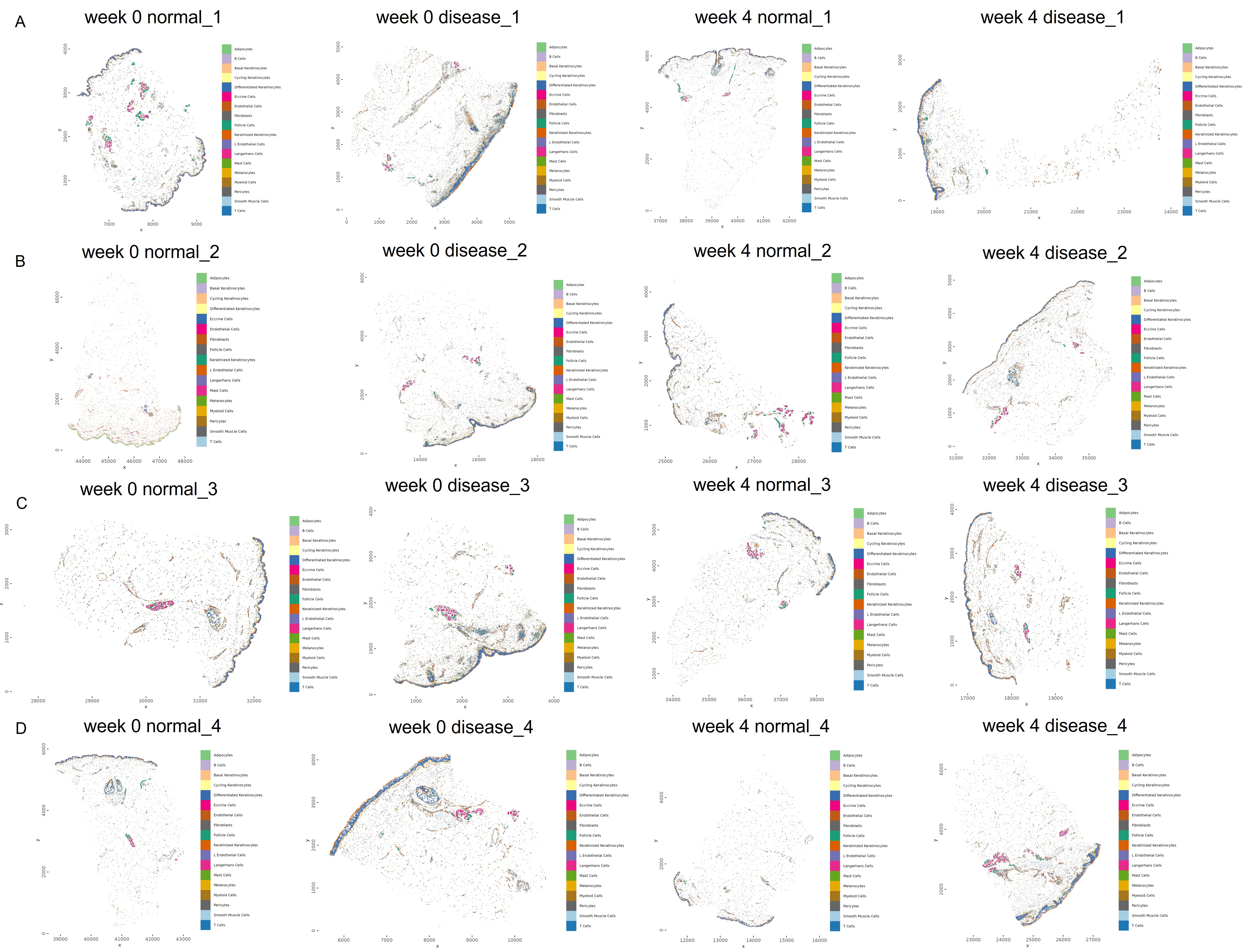

**Lesional Baseline**

**Lesional Week 4**

**Patient 1**

**Patient 2**

**Patient 3**

**Patient 4**

**A.**

**B.**

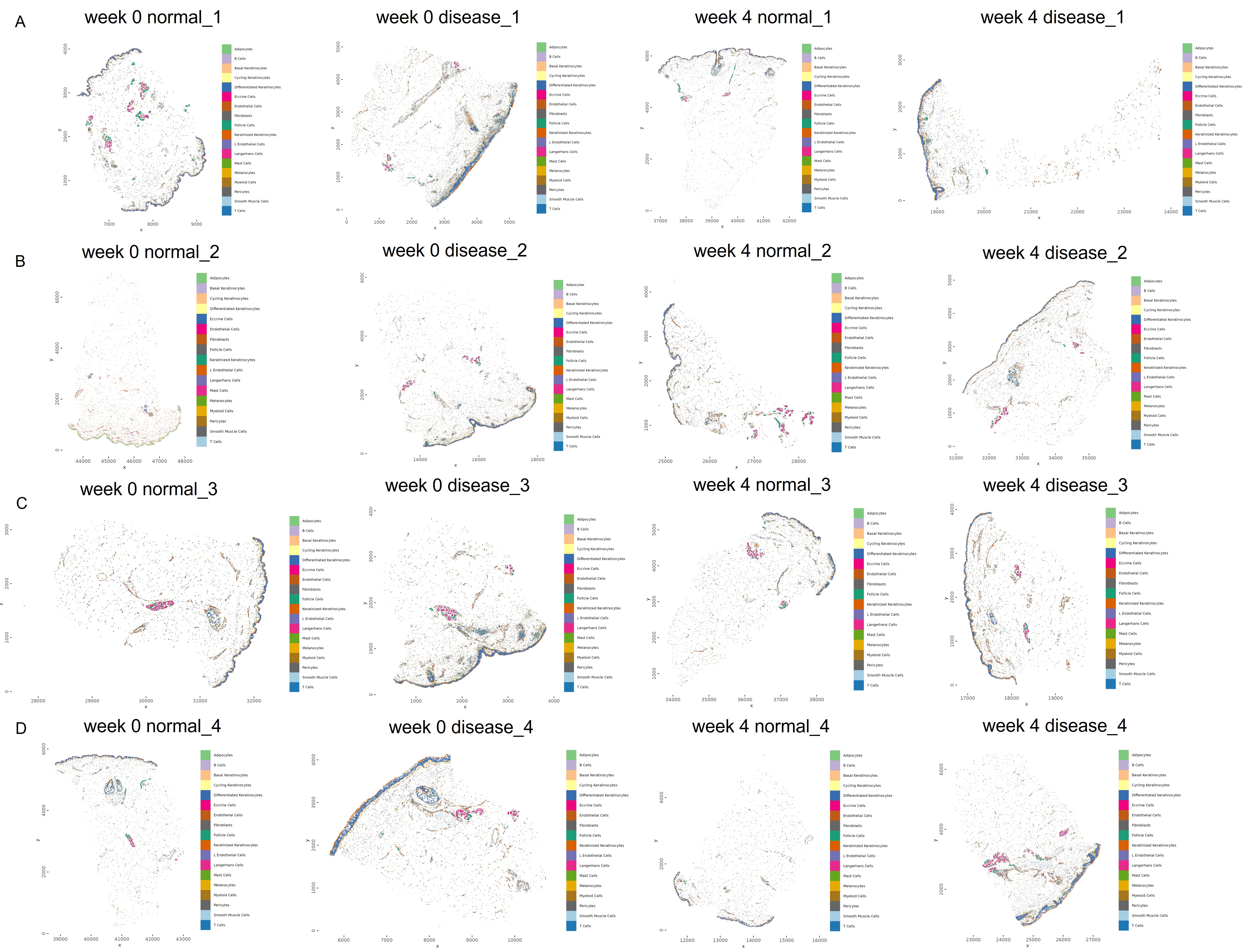

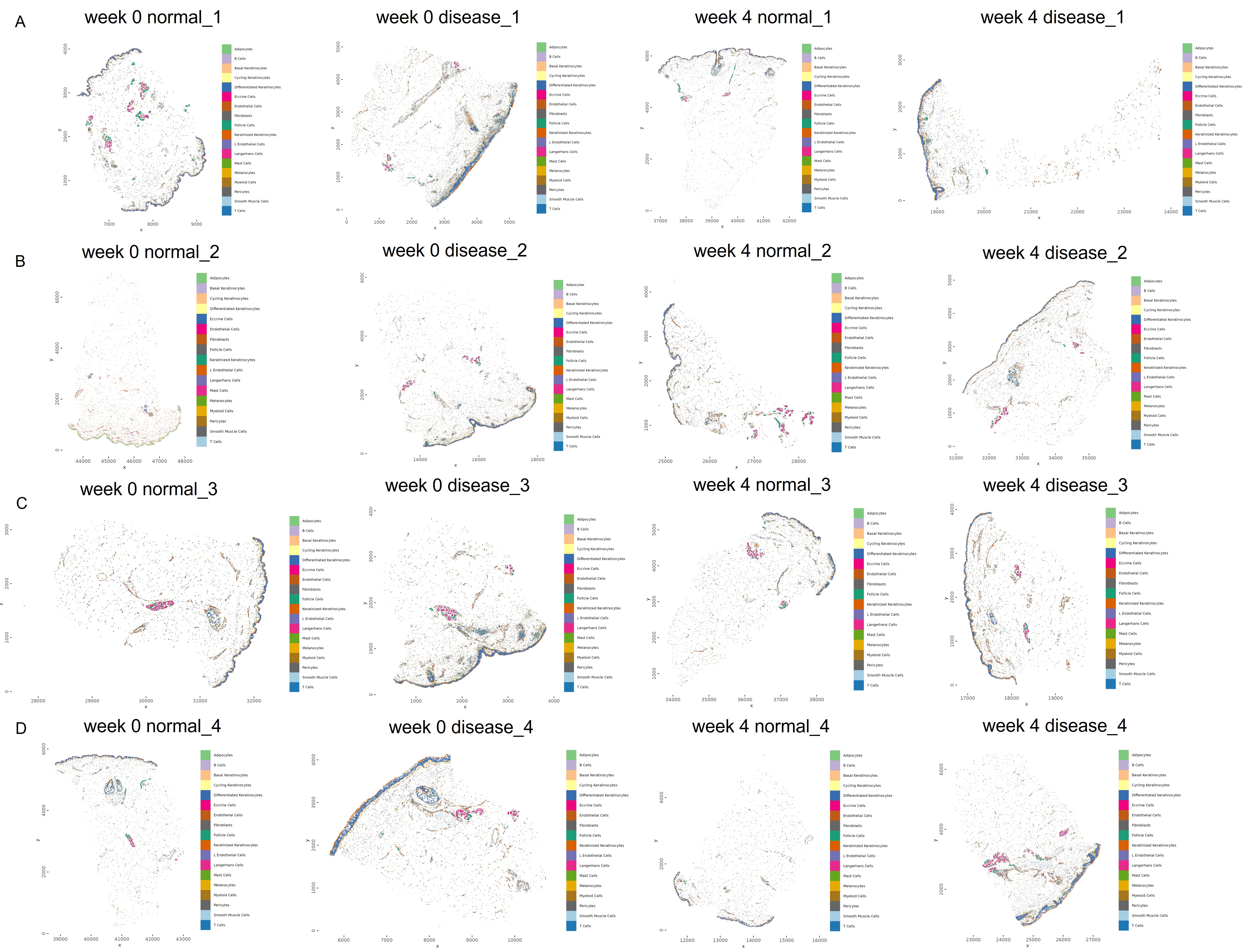

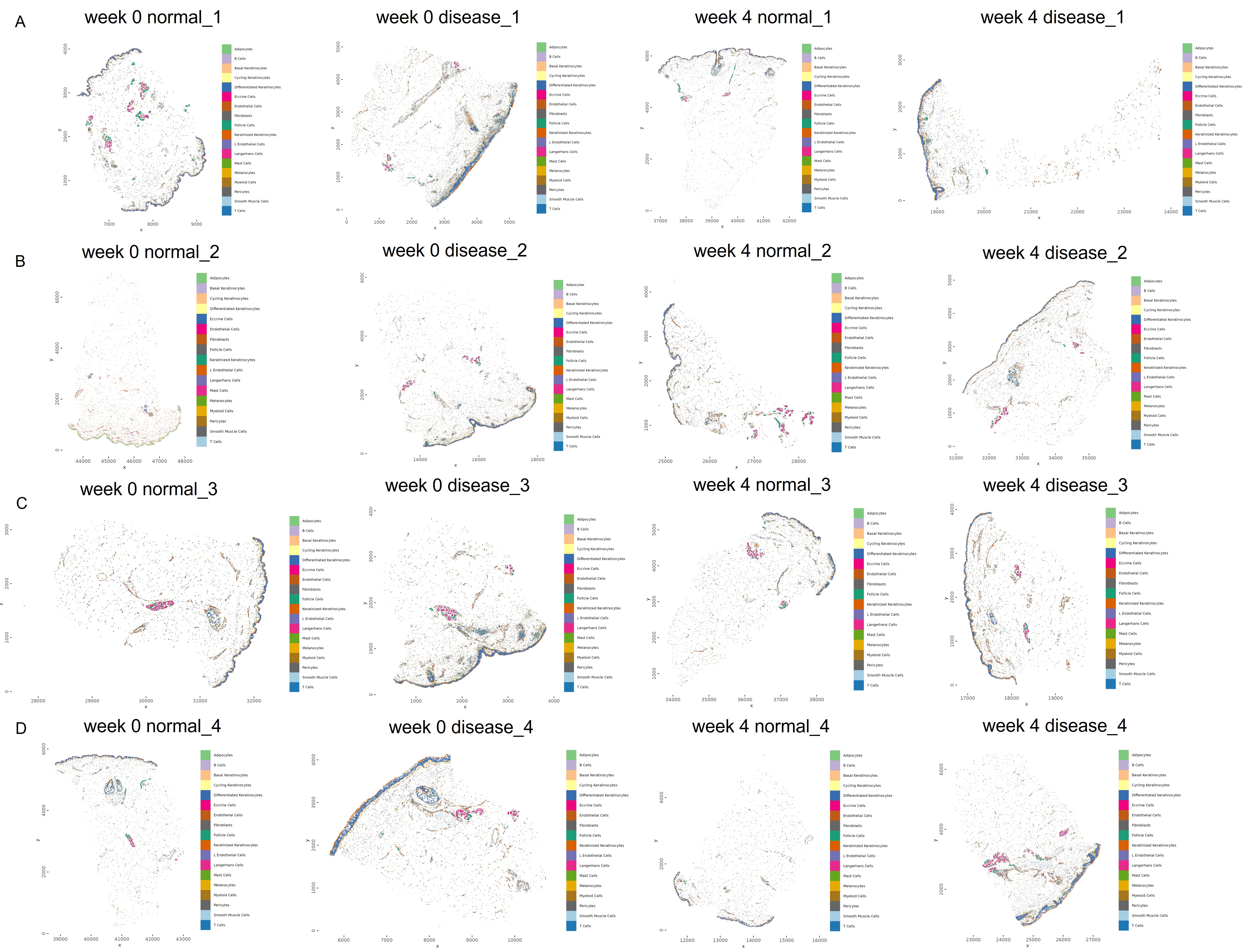

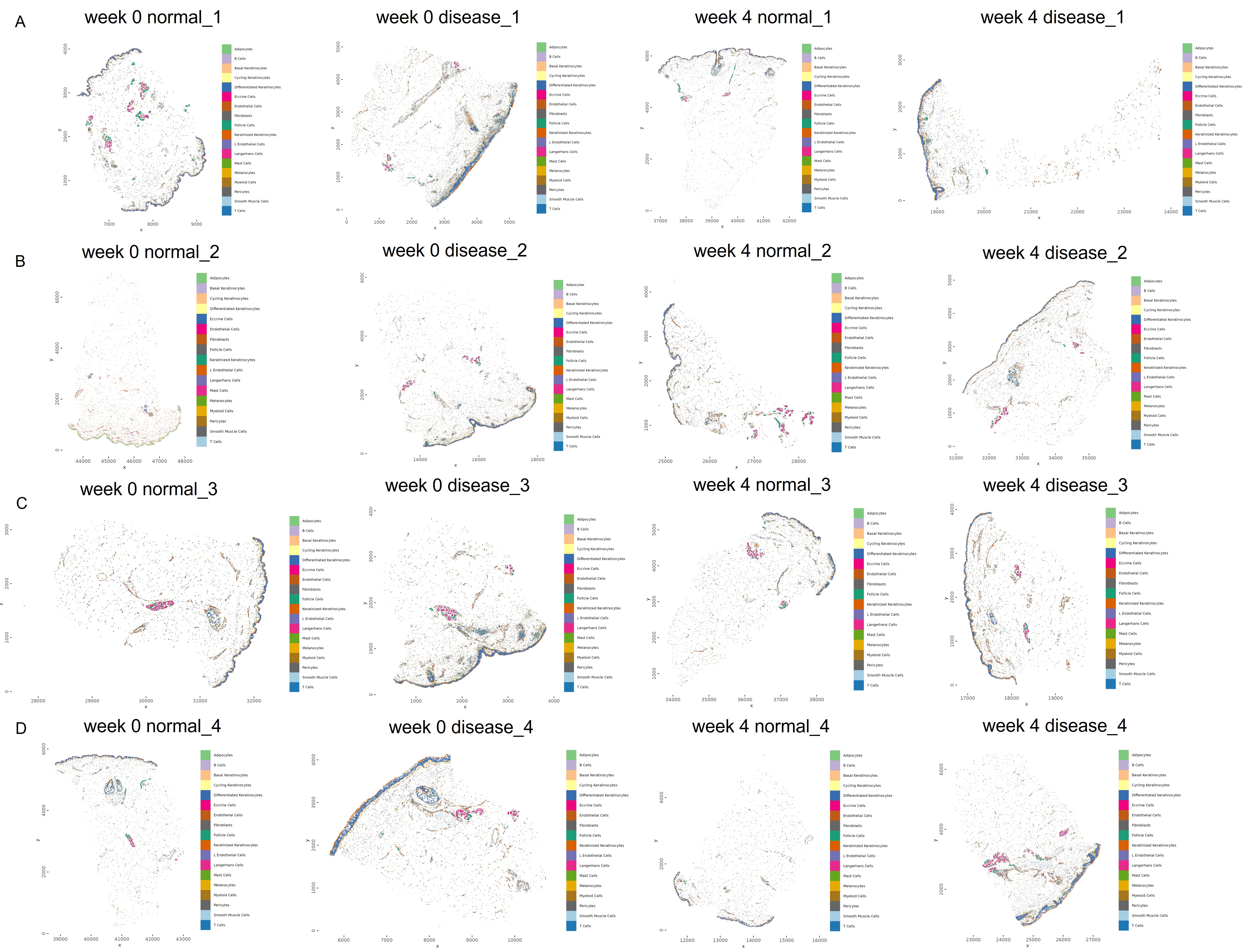

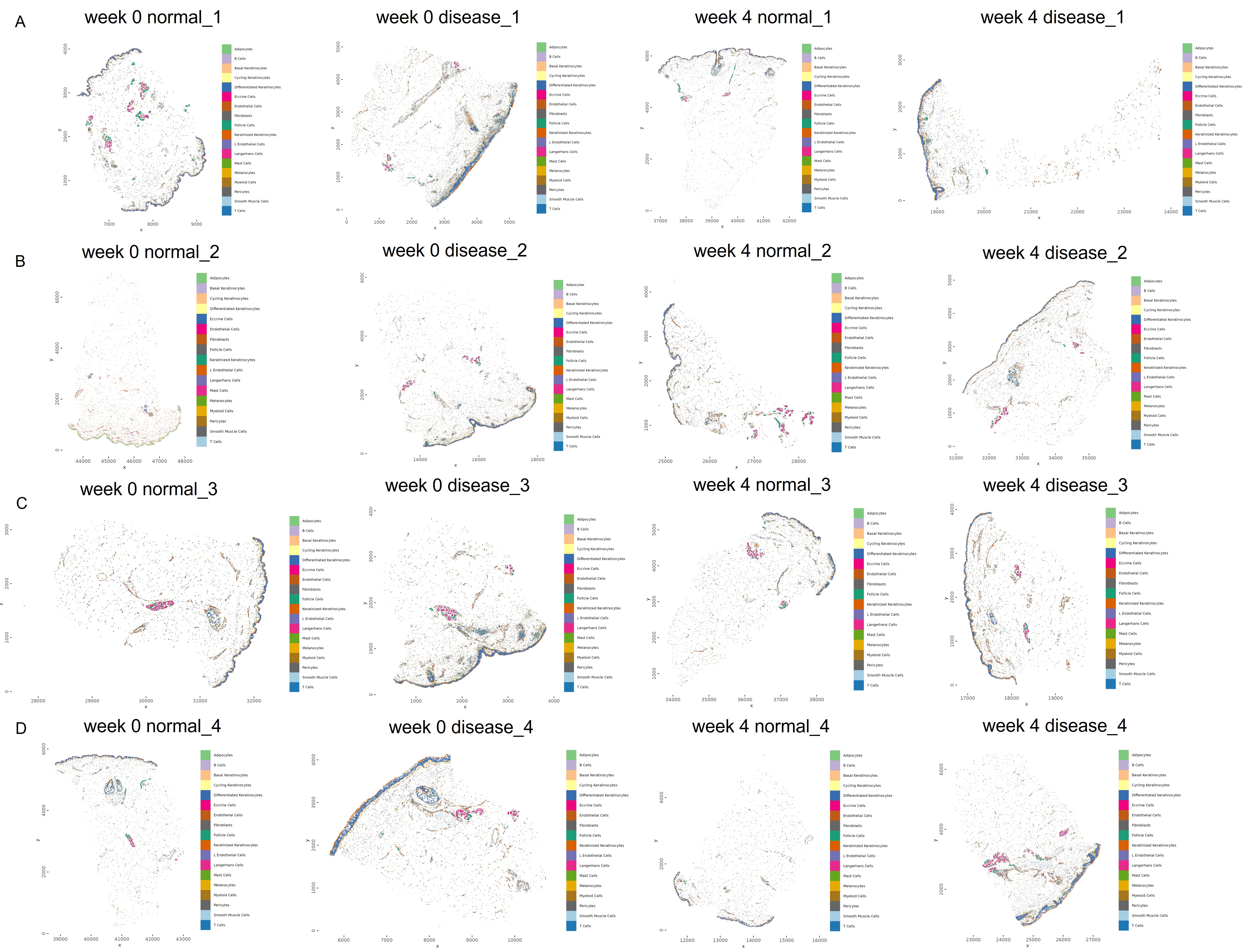

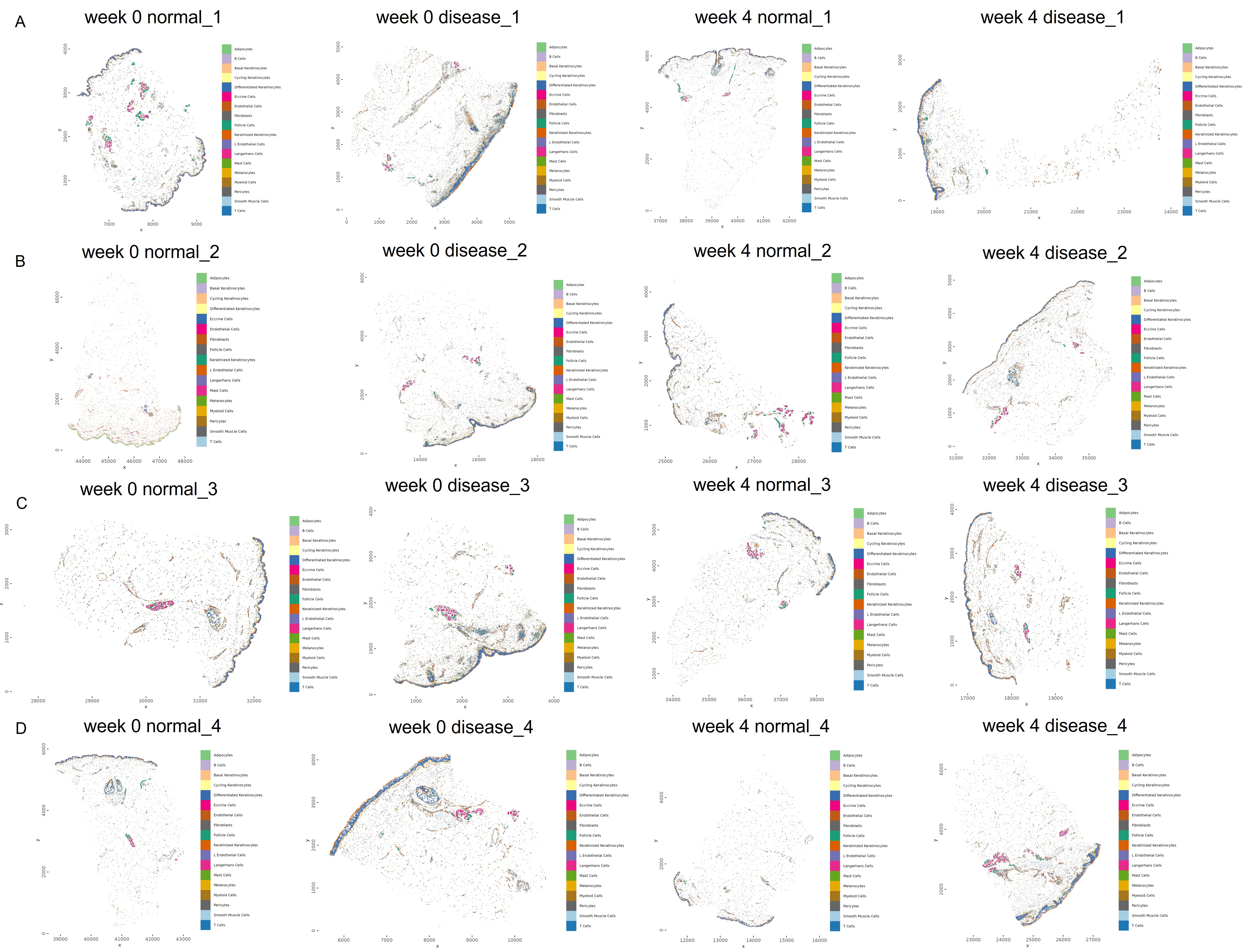

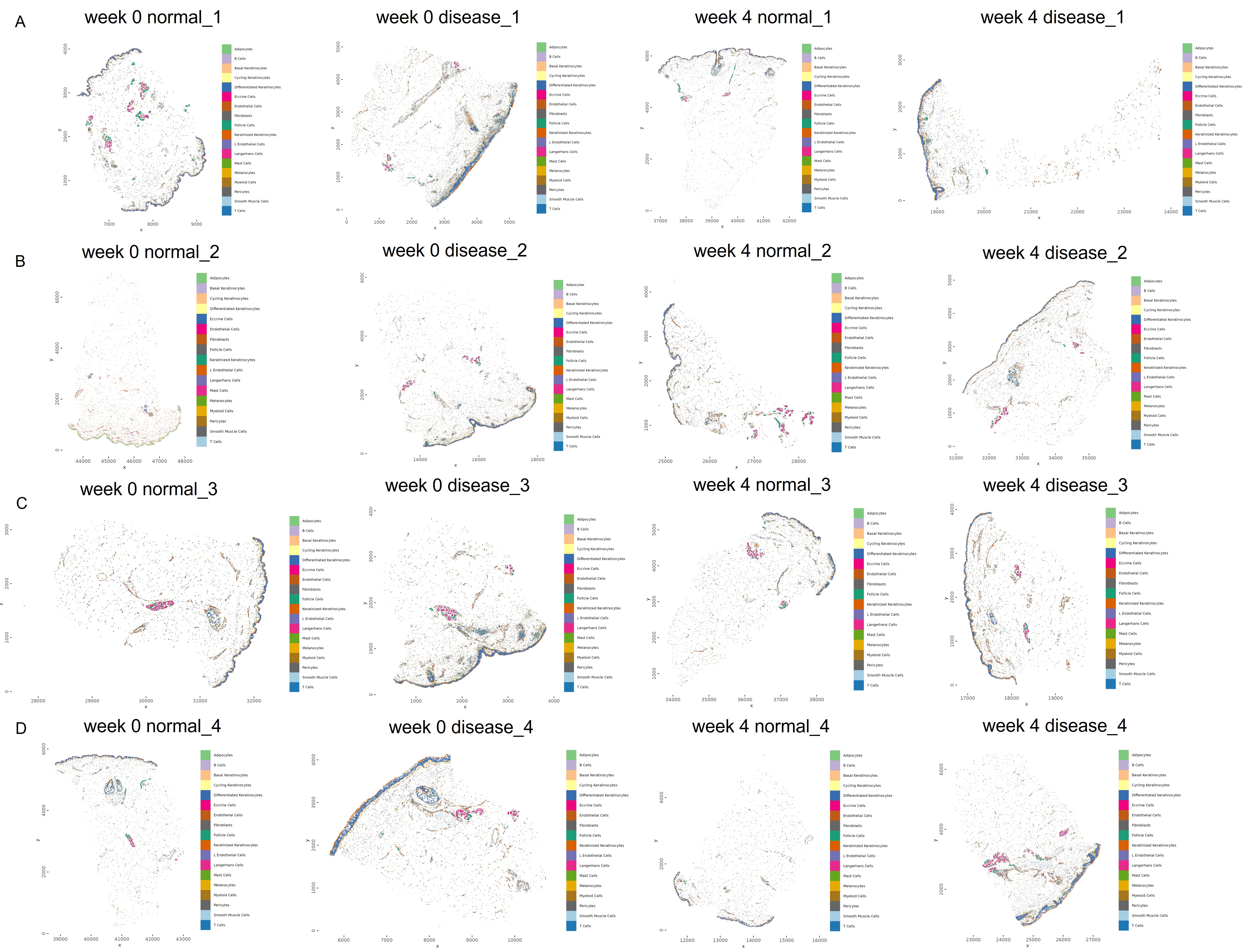

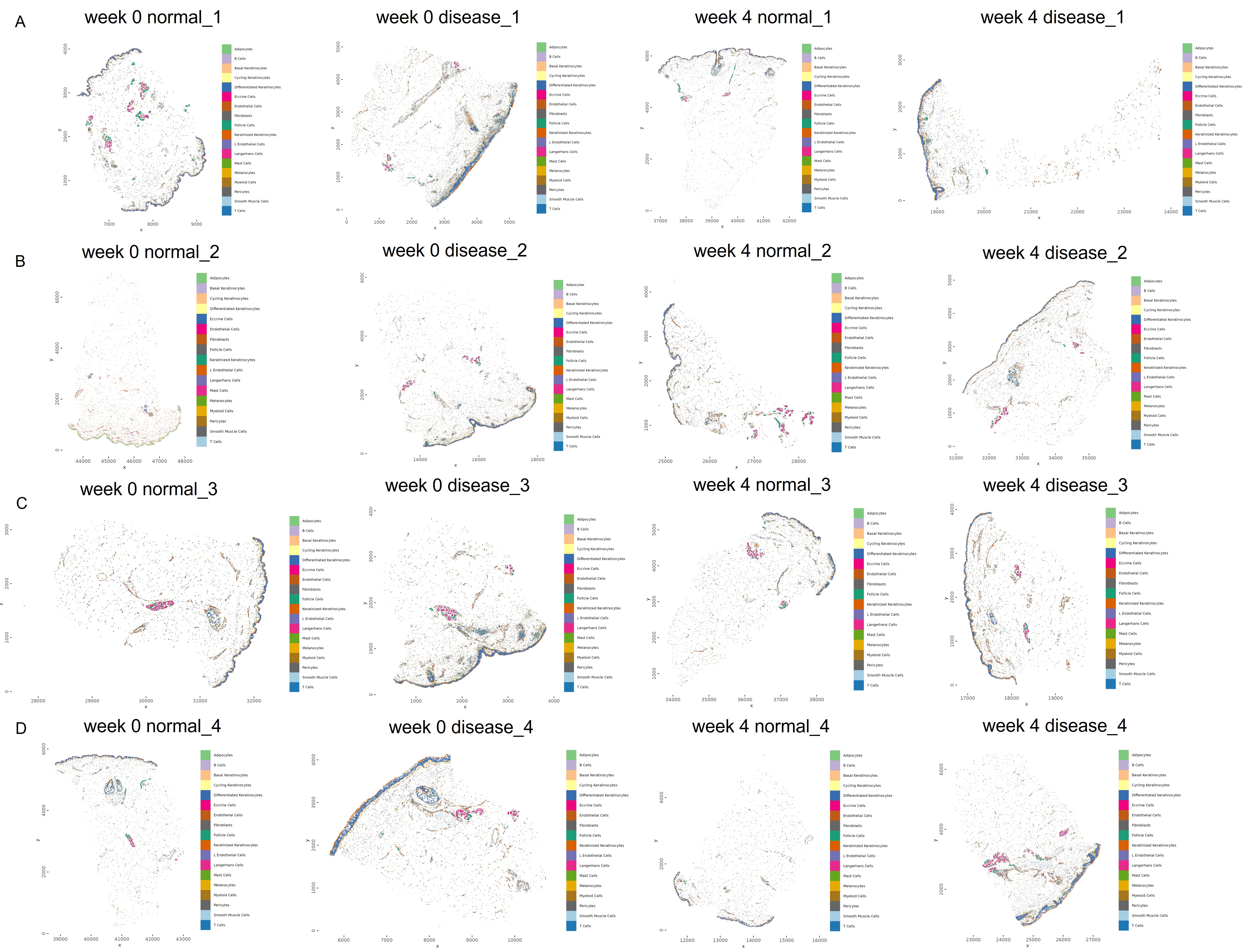

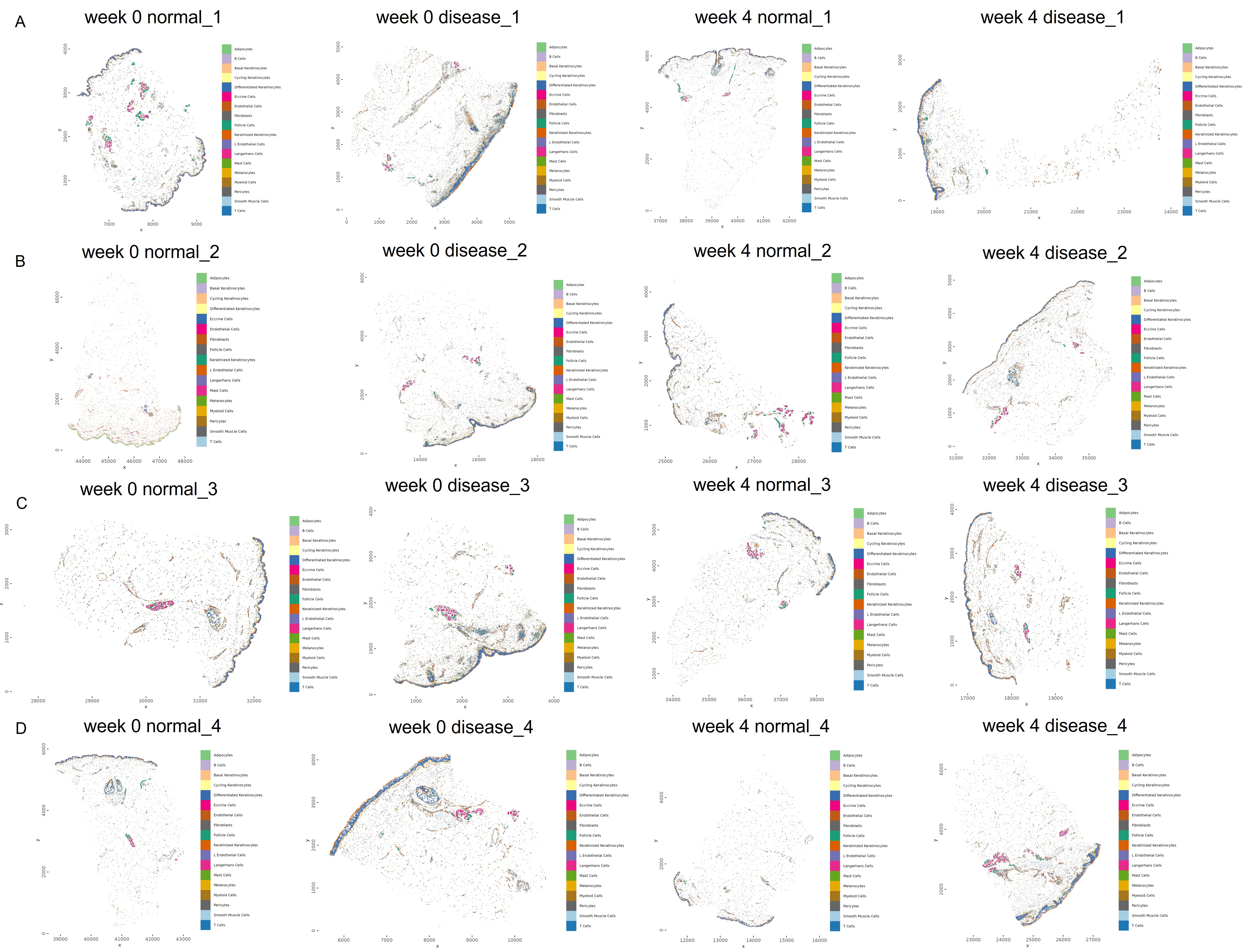

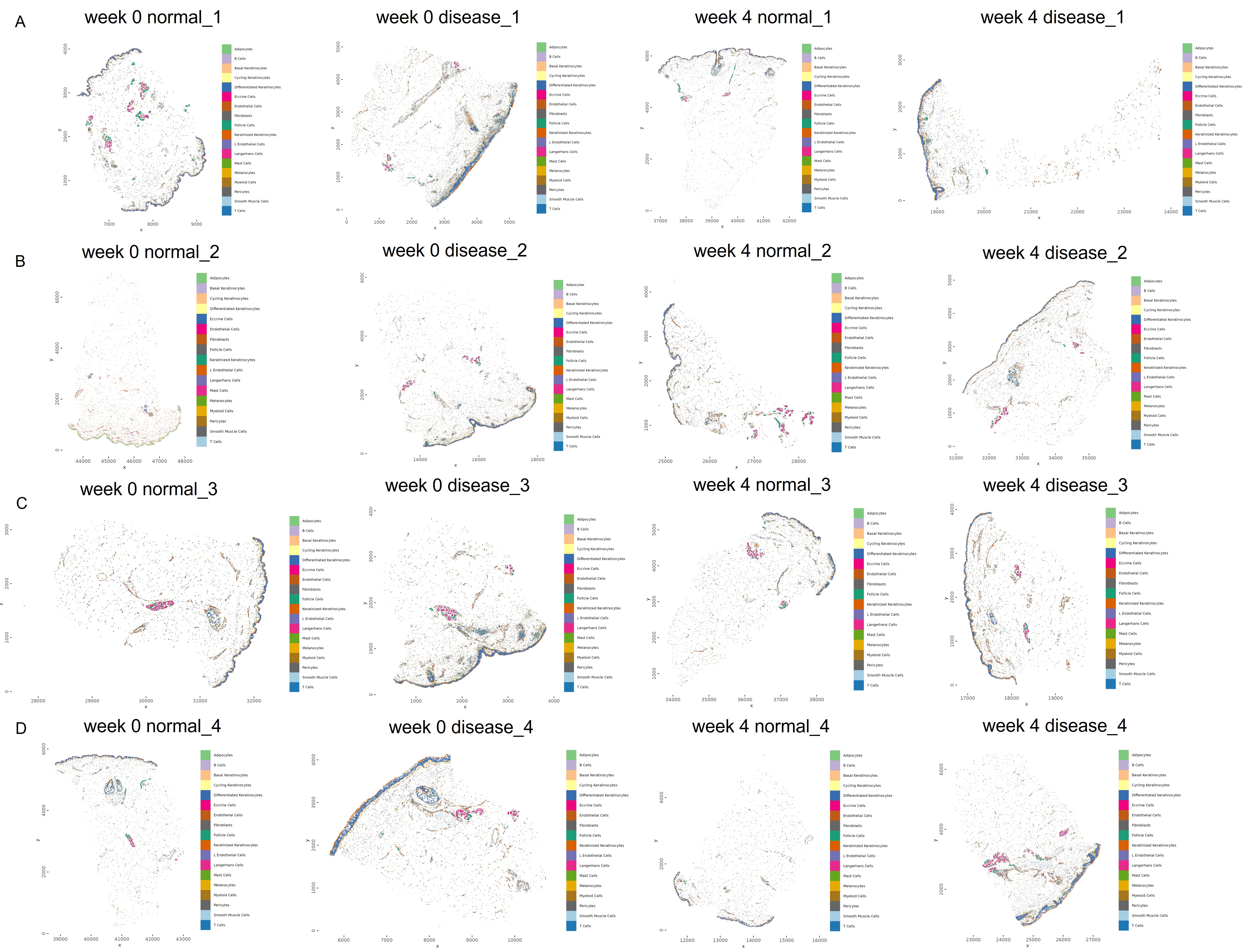

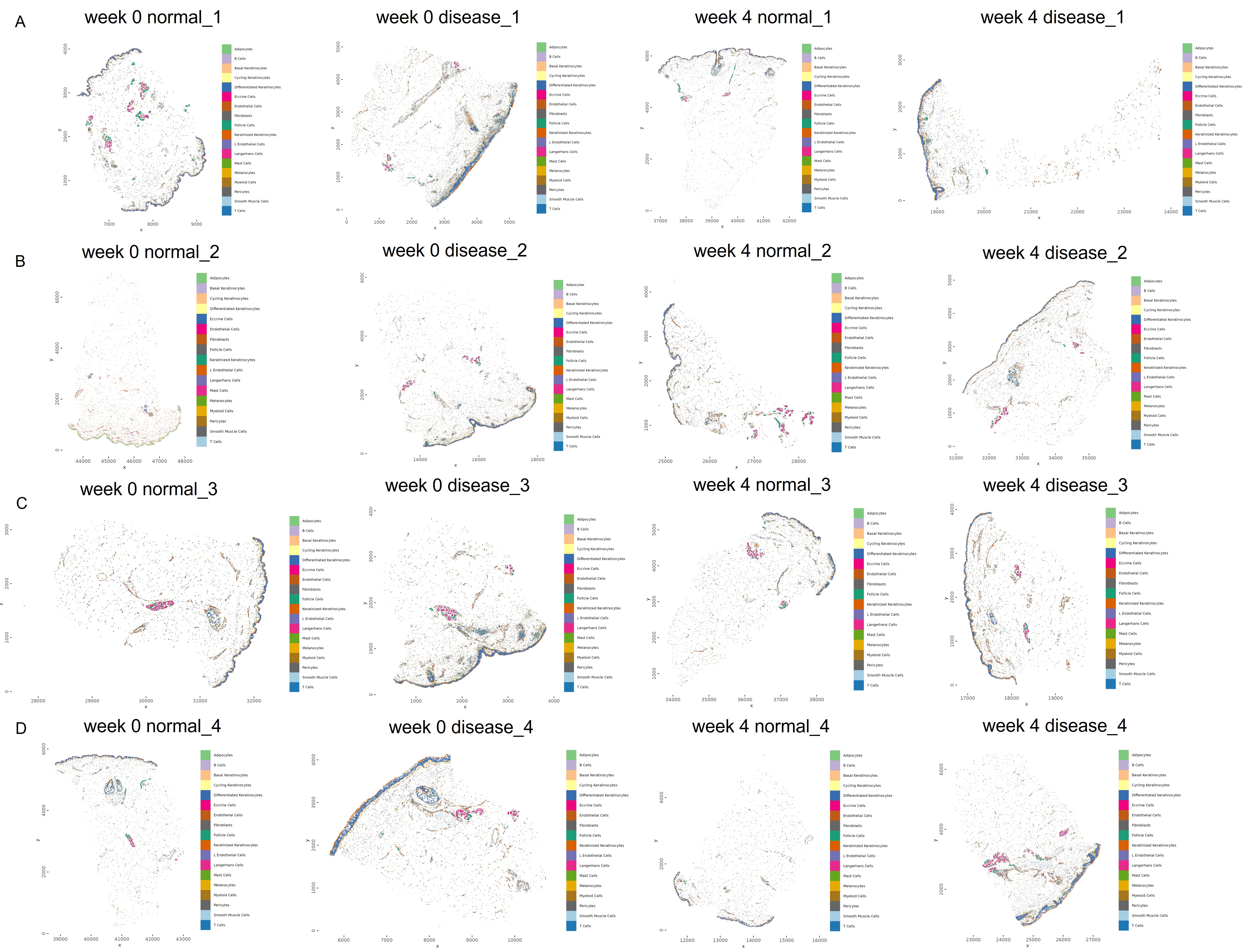

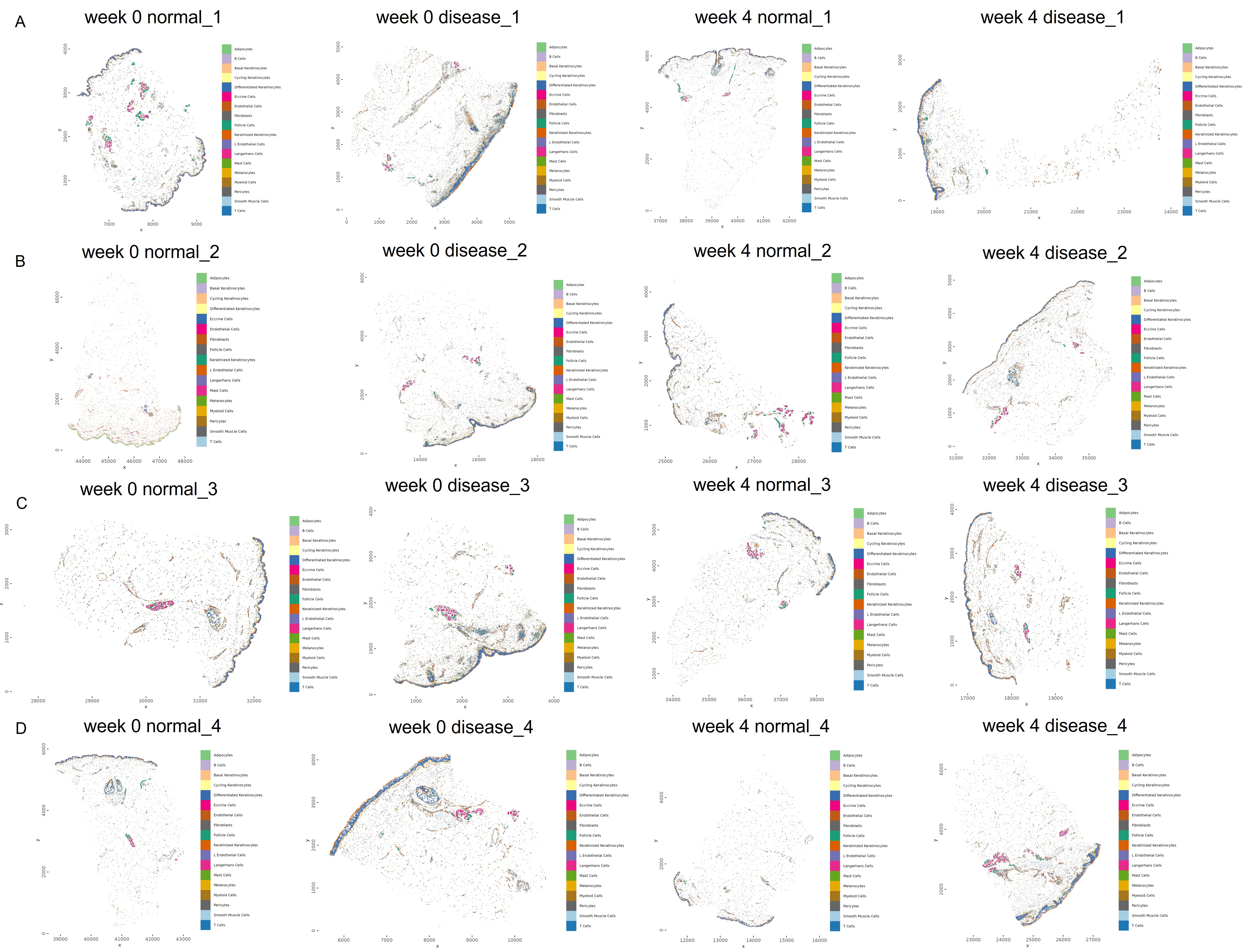

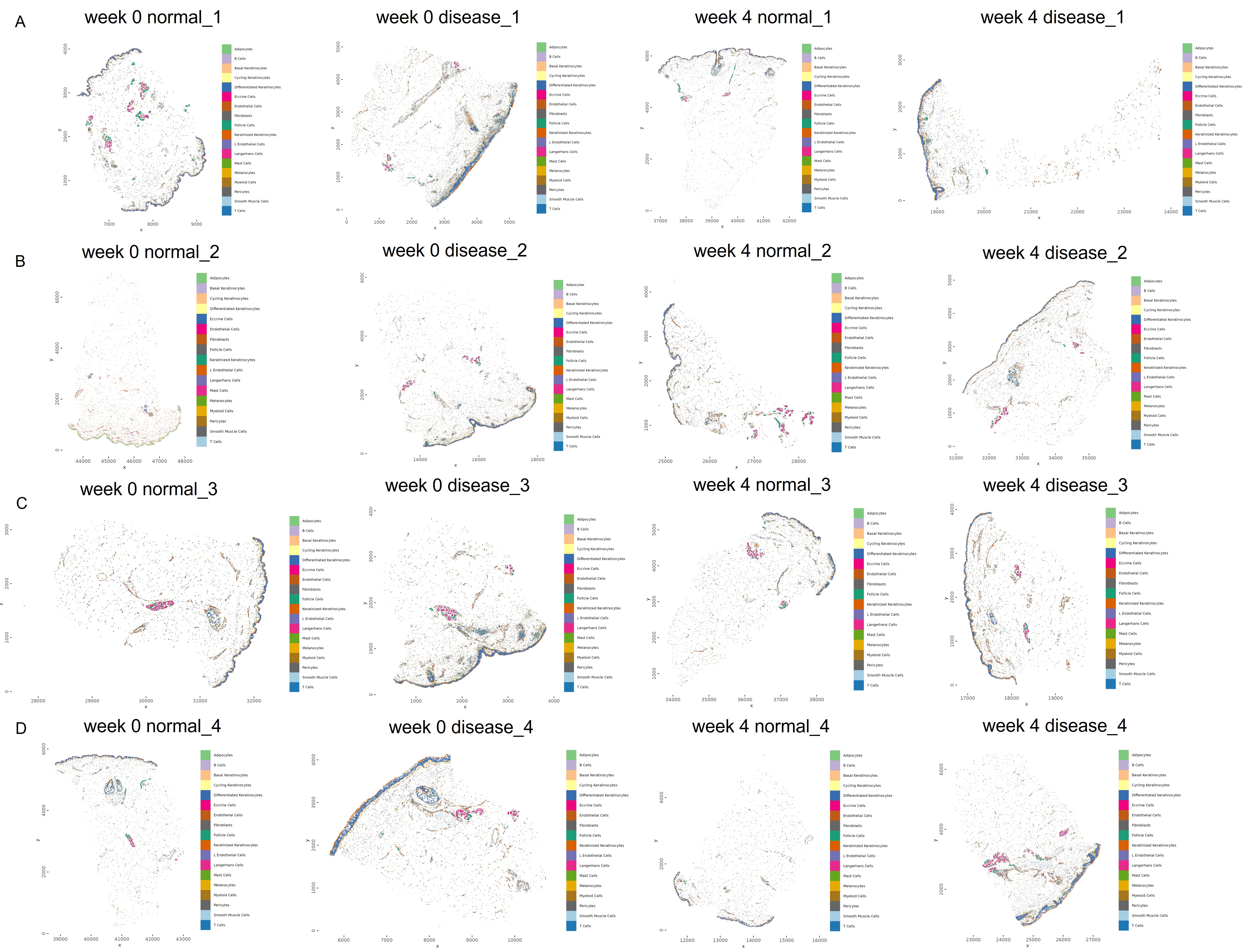

**Figure S4: Characterization of Molecular Responses to Brepocitinib in T-cells**

A. UMAP plot showing 6,679 T-cells from lesional dermatomyositis (DM) skin, colored by annotated T-cell subtype.

B. UMAP visualization displaying the number of differentially expressed genes (DEGs) per T-cell cluster at week 4 compared to baseline, highlighting heterogeneity in transcriptional responsiveness across T-cell subsets.

C. Gene Ontology (GO) pathway enrichment analysis of downregulated genes in CD8^+^ Tem cells at week 4 compared to baseline highlights decreased expression of pathways related to antiviral and cytotoxic responses.

D. Violin plots showing reduced expression of cytotoxic effector genes in CD8^+^ Tem cells following 4 weeks of brepocitinib treatment.

E. Boxplots illustrating individual cell AUC scores for Type I interferon (IFN), Type II IFN, and IL2-STAT5 signaling activity across T-cell subsets.

F. Volcano plot showing DEGs in NK cells at week 4 compared to baseline, demonstrating downregulation of IFN-stimulated genes.

G. GO pathway analysis of downregulated genes in NK cells at week 4 compared to baseline, revealing suppression of antiviral defense and viral response pathways.

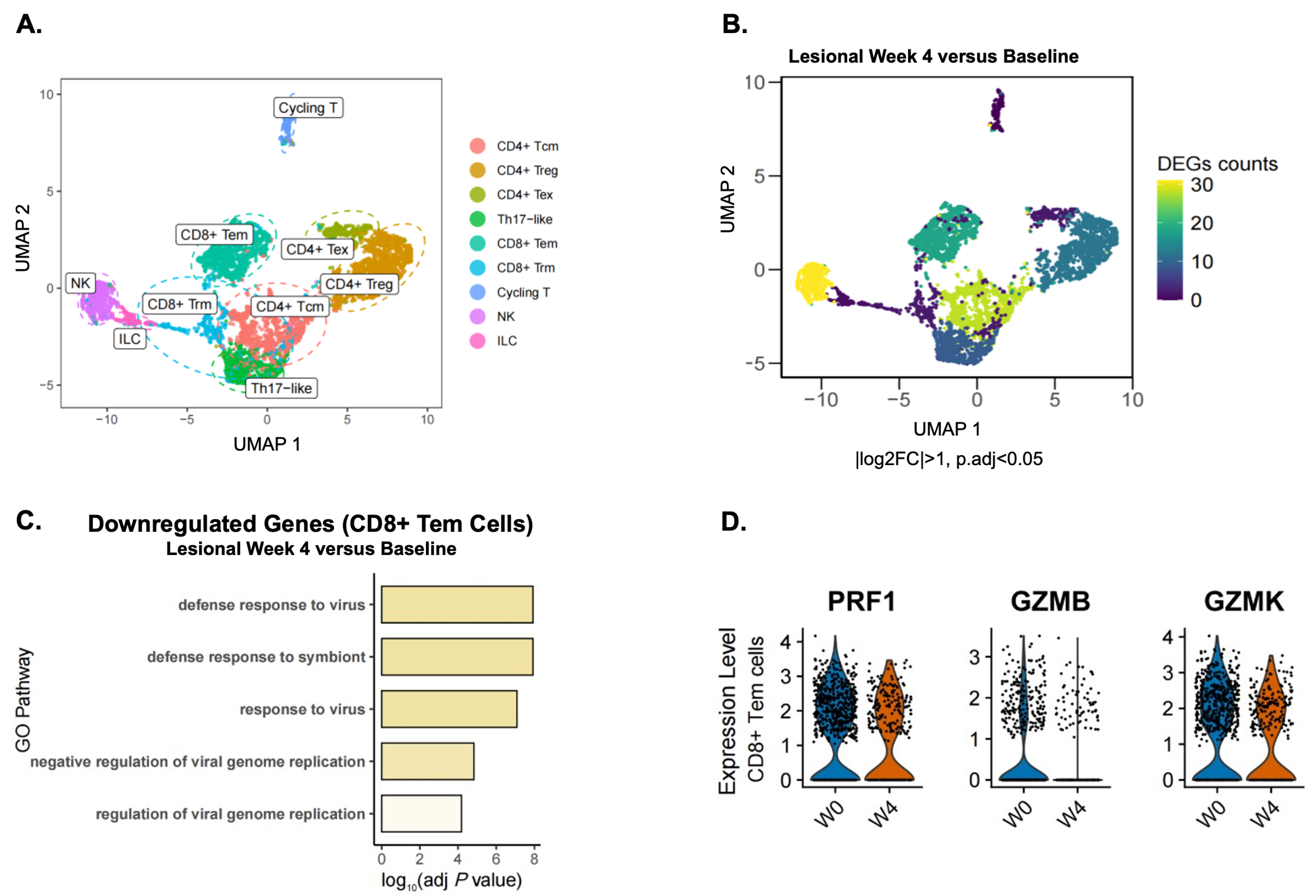

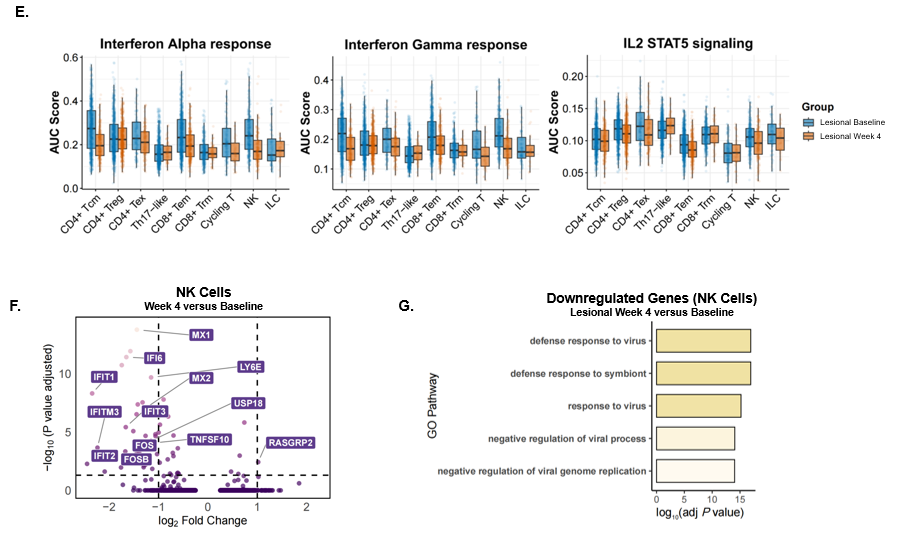

**Fig. S5: Characterization of Molecular Responses to Brepocitinib in Keratinocytes**

A. Dot plot displaying differentially expressed genes (DEGs) across keratinocyte subtypes (basal, cycling, differentiated, and keratinized keratinocytes) at week 4 compared to baseline in lesional DM skin.
B. Gene ontology (GO) pathway enrichment analysis of downregulated genes in cycling (left) and basal (right) keratinocytes at week 4 compared to baseline, highlighting suppression of antiviral defense, viral response, and type I IFN-related pathways.
C. Chord diagram illustrating altered intercellular communication between cycling keratinocytes and T-cell subpopulations at week 4 compared to baseline.

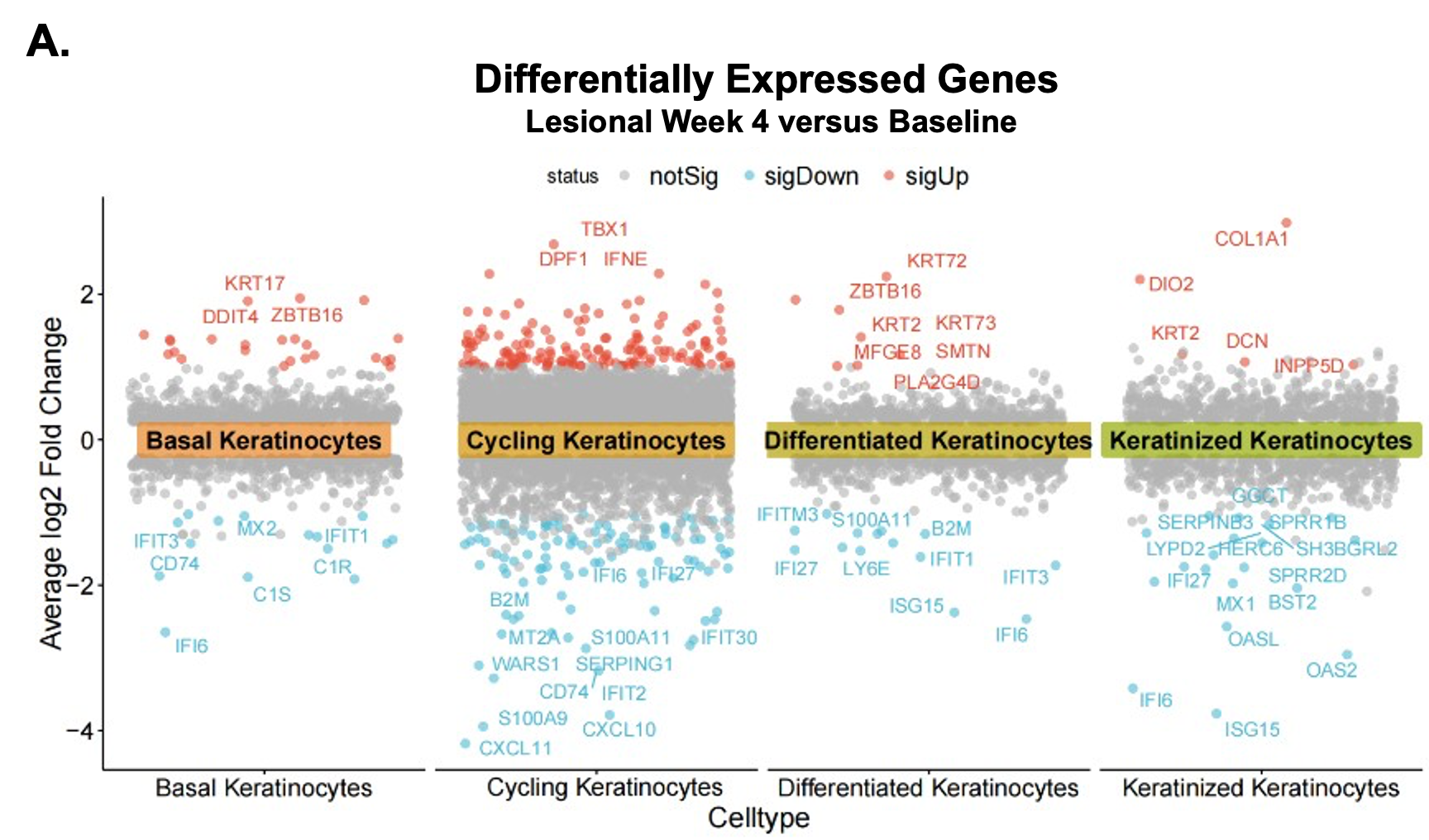

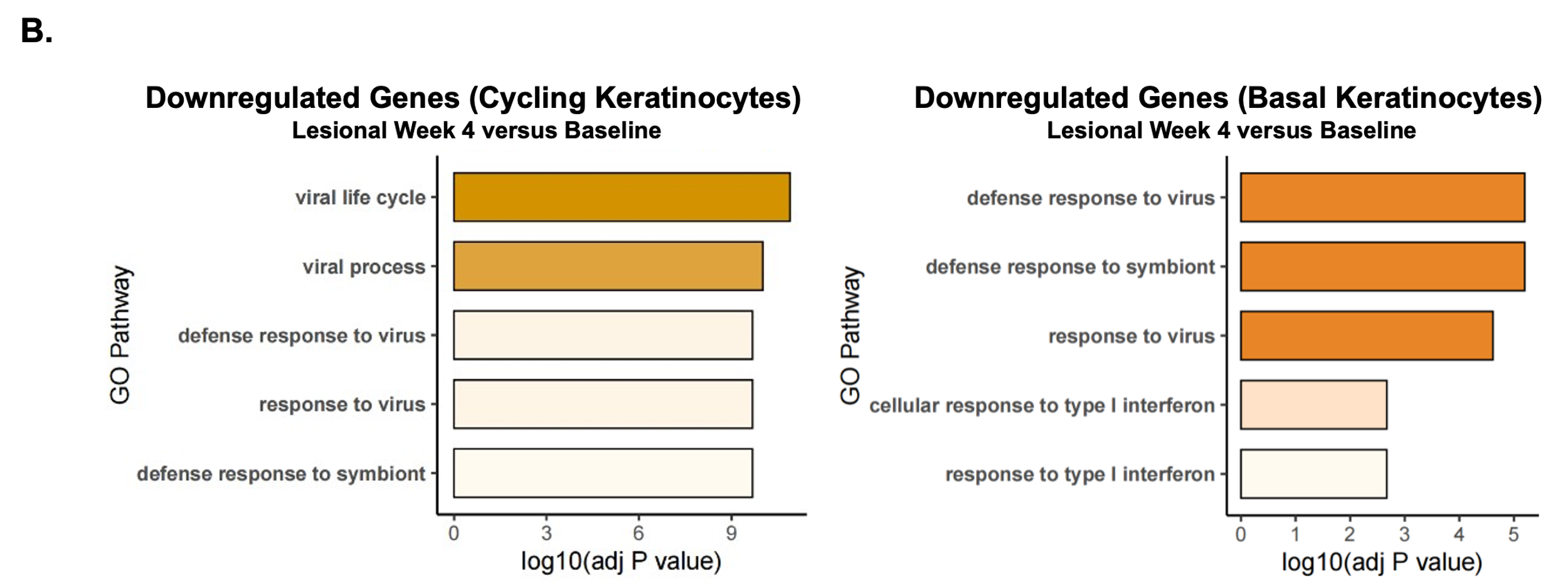

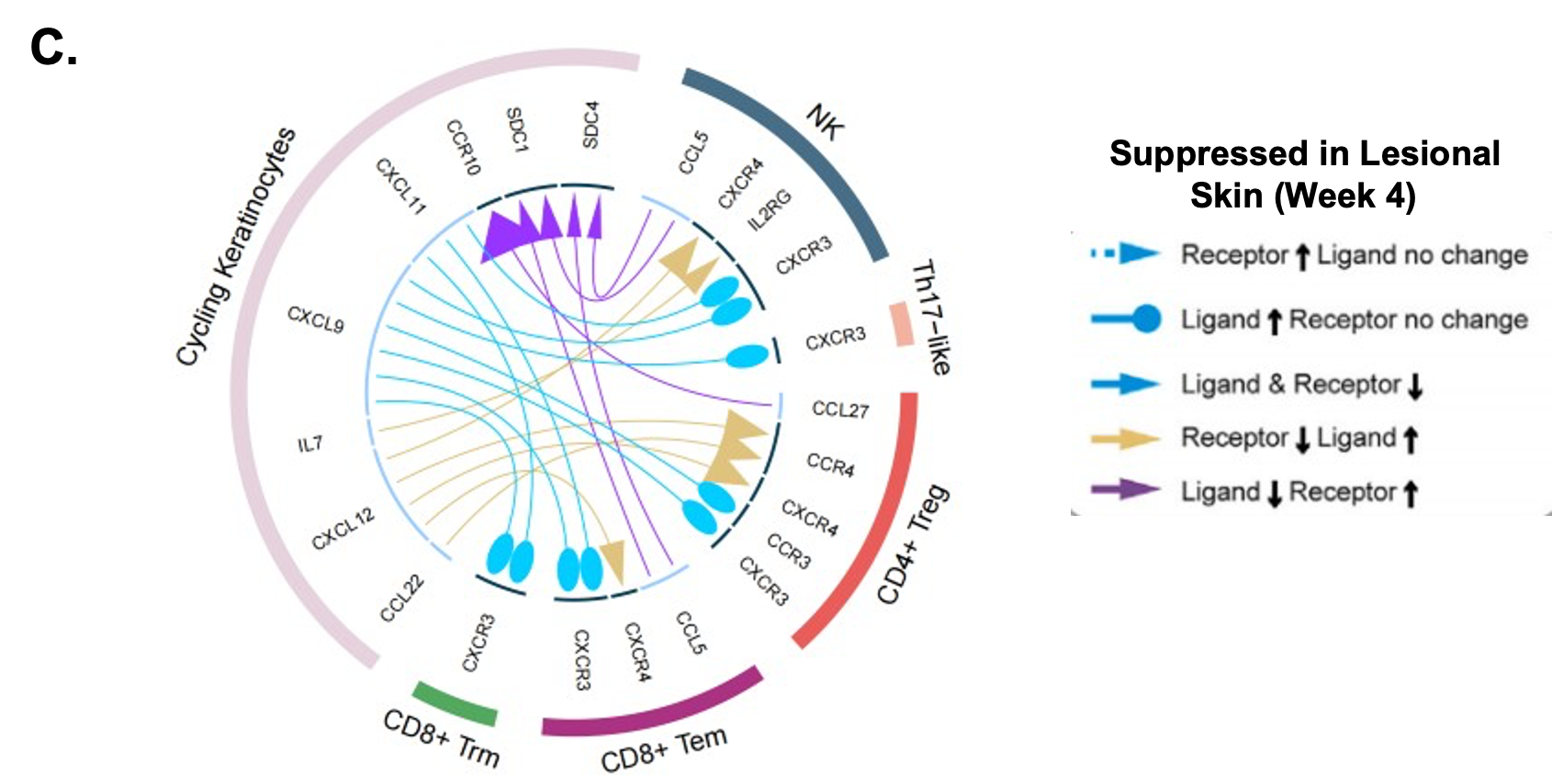

**Figure S6: Characterization of Molecular Responses to Brepocitinib in Pericytes**

A. Gene set enrichment analysis (GSEA) at week 4 versus baseline in pericytes highlights suppression of pathways related to extracellular matrix remodeling, angiogenesis, and cellular migration.
B. Volcano plot showing differentially expressed genes (DEGs) in pericytes at week 4 compared to baseline illustrates broad downregulation of inflammatory, matrix-associated, and migratory gene programs following brepocitinib treatment.
C. Gene ontology (GO) pathway analysis of downregulated genes in pericytes at week 4 versus baseline demonstrates decreased activity in pathways related to extracellular matrix organization, epithelial/vascular cell migration, and structural remodeling.
D. Predicted ligand-target regulatory matrix showing the top-ranked ligands identified from intercellular signaling analyses and their inferred gene targets in pericytes.
E. NicheNet analysis of potential ligands expressed by CD8⁺ Tem cells ranked by their correlation with observed pericyte gene expression changes between week 4 and baseline, identifying T-cell–derived signals most likely to regulate pericyte transcriptional responses.
F. Chord diagram demonstrating altered intercellular communication between pericytes and T-cell subpopulations at week 4 versus baseline, revealing reduced inflammatory crosstalk and attenuated ligand–receptor interactions with brepocitinib.

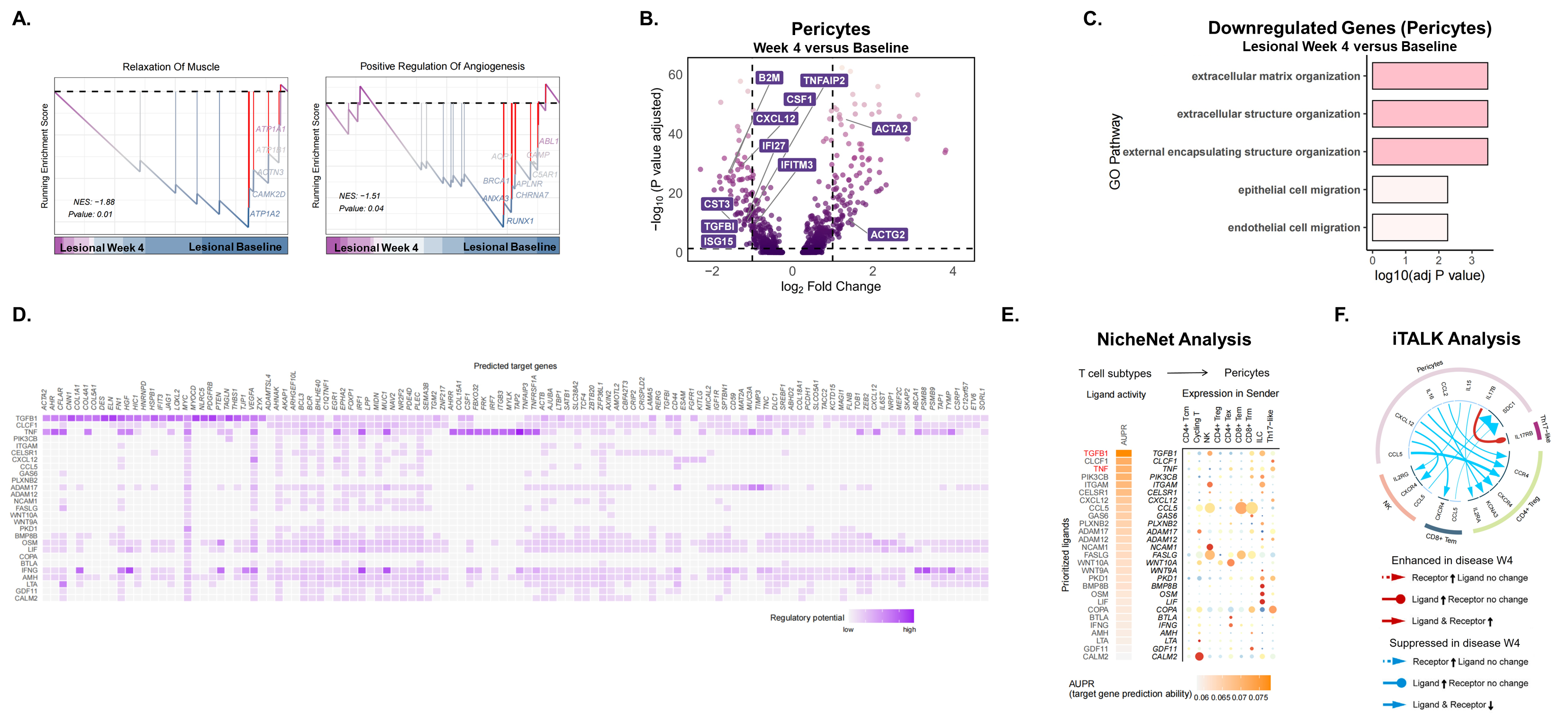

**Figure S7: Characterization of Molecular Responses to Brepocitinib in Myeloid Cells**

A. UMAP plot of myeloid cells from lesional DM skin, colored by annotated subtype.
B. UMAP visualization showing the number of differentially expressed genes (DEGs) per myeloid subcluster at week 4 compared to baseline.
C. Relative proportion of each myeloid cell subtype within the global single-cell dataset at baseline and week 4.
D. Volcano plot of DEGs in cDC2B cells comparing week 4 with baseline, demonstrating downregulation of interferon-stimulated genes and other inflammatory mediators.
E. Gene ontology (GO) pathway analysis of DEGs in cDC2B cells, highlighting suppression of antiviral defense, viral response, and negative regulation of viral genome replication pathways at week 4 compared to baseline.

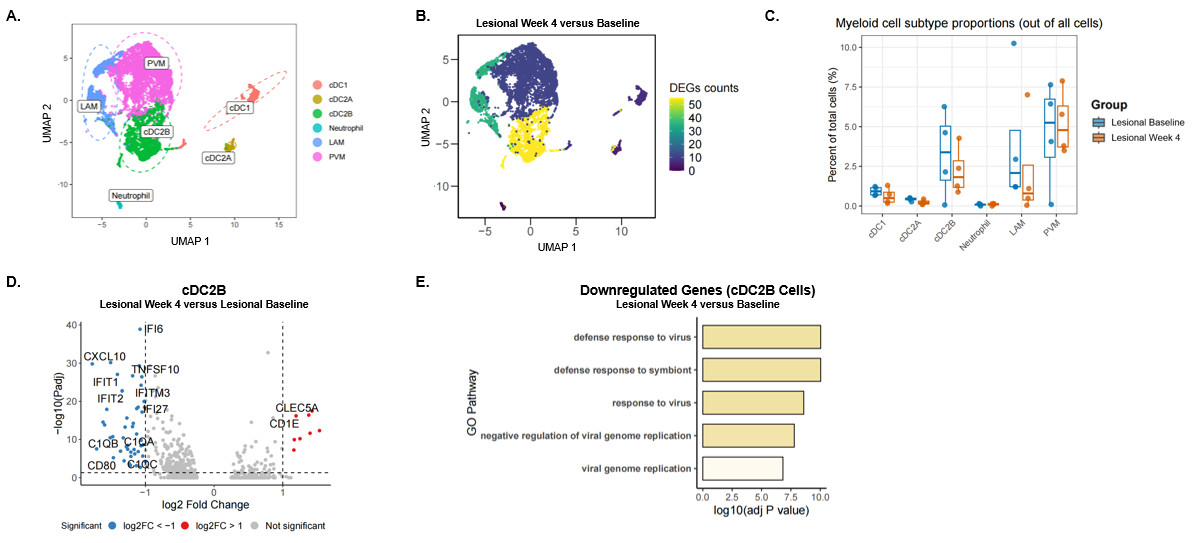
